# Estimating the impact of different Intermittent Preventive Treatment in Pregnancy delivery strategies on low birth weight outcomes under moderate and high malaria transmission settings: A modelling study

**DOI:** 10.64898/2026.05.30.26354497

**Authors:** Levoniah Chakuvinga, Caroline Franco, Sheetal Silal

**Affiliations:** Modelling and Simulation Hub Africa, Department of Statistical Sciences, University of Cape Town, Cape Town, South Africa; Biostatistics and Health Data Science, Institute of Applied Health Sciences, School of Medicine, Medical Sciences and Nutrition, University of Aberdeen, King’s College, Aberdeen AB24 3FX, United Kingdom; Centre for Global Health, Nuffield Department of Medicine, Oxford University, Oxford, United Kingdom

## Abstract

**Introduction:** Malaria during pregnancy is a major risk factor for low birth weight (LBW) in newborns, which in turn negatively affects the growth and development of the child. The World Health Organization (WHO) recommended interventions for pregnant women living in malaria endemic countries that include the use of intermittent preventive treatment in pregnancy (IPTp). However, WHO asserts that the coverage of pregnant women taking the recommended doses of IPTp are still very low. The primary goal of this study was to estimate the effects of increasing the coverage of doses of IPTp and to assess the effect of pregnancy timing in relation to seasonal transmission on malaria infections during pregnancy and neonates with LBW. We explored these effects in moderate and high transmission settings.

**Methods and Findings:** A compartmental mathematical model depicting malaria during pregnancy with IPTp doses was formulated to analyze the effects of IPTp, insecticide treated net (ITN) use and seasonal variations in moderate and high malaria transmission settings. Our simulation findings suggest that increasing both ITN use and IPTp dose coverages to high levels, prevents 90% and 84% clinical cases for pregnancies starting in August in moderate and high transmission, respectively. Our model predicts that increasing the coverage of the first dose of IPTp to 90%, while lowering subsequent doses, averts 44% and 37% LBW cases for the August cohort in moderate and high transmission settings, respectively. Unprotected pregnancies overlapping the January peak in rainfall and malaria incidence during the third trimester experience the highest LBW burden.

**Conclusions:** The highest IPTp coverage prevents the highest number of LBWs providing evidence of the benefits of scaling up IPTp. Overall, our results demonstrate that increasing ITN use has a substantial impact in reducing clinical malaria cases during pregnancy and improves birth outcomes. This highlights its importance as a key intervention, and the health benefits it would provide for malaria control goals for pregnant women. Pregnancies that overlap with the epidemic peaks in later trimesters lead to a rise in LBWs, indicating the necessity of protecting pregnant women at risk of malaria infection till delivery.

## Introduction

Malaria in pregnancy (MiP) is a major global cause of maternal morbidity, leading to stillbirth, fetal anemia, low birth weight (LBW) for the newborn and premature delivery and anemia for the mother [1, 2]. There were an estimated 12.5 million cases of MiP in the World Health Organization-Africa (WHO-AFRO) region in 2023, and the southern and eastern Africa region contributed 26.3% of the cases, which was a major improvement of 564,000 MiP infections from the previous two years [2–4]. LBWs defined as babies born weighing less than 2500g despite gestational age, remain a global health concern and WHO has a target to achieve 30% reduction in LBWs by 2030 [5].

According to the World Malaria Report of 2025, there are still about 411,000 LBWs in the African region due to MiP that could be averted and this could be achieved by increasing the coverage of IPTp [3]. A significant rise in newborn mortality and long-term illness is linked to LBWs and thus prevention of a significant number of LBWs will save lives [5, 6].

The WHO recommendations for malaria prevention and control during pregnancy in areas of stable malaria transmission include providing insecticide-treated nets (ITNs), intermittent preventive treatment in pregnancy (IPTp), and effective management of malaria and anaemia [7]. IPTp entails administering a full therapeutic dose of a strong antimalarial drug, sulfadoxine-pyrimethamine (SP), to all pregnant women whether infected by malaria or not and irrespective of the number of previous pregnancies [2, 8]. Despite growing levels of resistance to SP in a few parts of Sub-Saharan Africa, IPTp-SP is likely to remain an effective intervention for preventing malaria-related LBW because it has proved to effectively prevent malaria infections and related LBWs [9]. The WHO advises that this preventive treatment be given at each monthly prenatal visit, ideally beginning in the early stages of the second trimester of pregnancy until delivery [7].

Pregnant women who receive only one or two dose(s) of IPTp are not fully protected and may contract malaria and risk having negative consequences in their pregnancy and the foetus. A diagrammatic representation of how IPTp is administered in countries like Zimbabwe is shown in Fig. S1 in the supplementary file. Zimbabwe for example, is currently implementing IPTp in 26 high malaria burden districts and the country aims to achieve a target of at least 85% of pregnant women getting more than three doses by 2026 [10]. However, the coverage of pregnant women receiving the recommended doses remains very low throughout the WHO African region as revealed by the World malaria reports [3, 4, 11, 12].

Previous research has highlighted the effects of IPTp on the adverse outcomes of MiP, supporting the WHO recommendations for MiP. Desai et al. (2018) emphasized that protecting pregnant women from malaria remains crucial even as overall transmission rates decrease [13]. Walker et al. (2014) pointed out that the most significant reductions in placental infection and LBW can be achieved by clearing or preventing infections early in pregnancy or just before conception. Additionally, using a mathematical model, Walker et al. (2017) assessed the potential impact of IPTp with sulfadoxine-pyrimethamine (IPTp-SP) by considering factors such as the inherent risk of malaria and the associated burden of LBW in the absence of protective measures [9, 14]. However, there appears to be very few studies which have looked at the population level of the impact of IPTp on pregnant women subpopulations [9, 14, 15].

Mathematical modelling involves the translation of a real-world problem into mathematics [16, 17]. It is valuable in exploring intricate phenomena, such as the population dynamics of infectious agents. From a population level perspective, mathematical models are useful tools that have been consistently used to simulate malaria dynamics incorporating seasonality and to evaluate the effectiveness of control interventions [17–19].

Malaria transmission usually follows seasonal patterns, with cases rising during and after the rainy season. Many mathematical models of malaria incorporating seasonality have been developed and analysed [9, 20–26]. Silal et al. (2014) included the seasonal nature of malaria transmission when they developed a mathematical model to assess the potential impacts of different interventions that may be used to achieve elimination in South Africa’s Mpumalanga district [26]. While previous models have tended to focus on assessing the effects of seasonality and a variety of interventions in general populations burdened with malaria, much attention was not paid to pregnant cohorts and, particularly, to LBW outcomes.

In this study we use mathematical modelling to represent the process of malaria during pregnancy for a cohort of women and incorporate IPTp and ITN interventions. In addition, we estimate the effects of increasing the coverage of IPTp doses on the number of malaria infections that occur in a cohort of pregnant women and the associated risk of neonates with LBWs in areas of moderate and high transmission intensity settings. We also use the model to analyse the impact of different pregnancy start times in the year and malaria seasonality on infections during pregnancy and the associated risk of neonates with LBW.

## Materials and methods

### Study setting

The simulation study was carried out for moderate malaria transmission and high malaria transmission settings. A moderate transmission area is defined as an area having an annual parasite incidence of 200-450 cases per 1000 population, and a high transmission area is defined as an area having more than 450 cases per 1000 population [27].

### Ethics

The study was cleared for ethical considerations by the University of Cape Town (UCT) research ethics committee (HREC REF 179/2024). Furthermore, the study’s protocol was reviewed and approved by the full board of the Medical Research Council of Zimbabwe (approval number: MRCZ/A/3363).

### Mathematical model for IPTp

We formulated a deterministic compartmental mathematical model of malaria in pregnancy with IPTp and ITNs as interventions drawing on the models of Cairns et al. (2011) [24] and Walker et al. (2013) [15]. The population in the model is a cohort of pregnant women who have access to ante-natal care (ANC). The cohort modelling approach was used in the model to track pregnant women for ten months, which is the maximum pregnancy period, as they go through the malaria transmission dynamics. At the end of ten months, we estimate the possible number of neonates born with malaria-related LBW from the cohort using the process described in the supplementary file. The mathematical model is a generalized susceptible-exposed-infected-recovered (SEIR) model with additional features for clinical and severe malaria. Infections in the model are classified as asymptomatic and clinical compartments. In the model, pregnant women begin in the susceptible state (S). Upon exposure to malaria infection, they enter the exposed state (E) where parasites are present in the bloodstream, but the women are not yet infectious. From there, women may progress to either the asymptomatic state (A) (infectious but without symptoms) or the clinical state (C) (infectious with clinical symptoms). The clinically ill women who do not receive timely treatment either transition to the severe state (*C_sev_*) or lose the clinical symptoms to become asymptomatic (A). Clinically ill and severe women can transition into treatment states (T and *T_sev_*), after which they lose infectiousness and move into the recovered state (R). Asymptomatic women recover naturally and enter the recovered state. In the recovered state, women have temporary immunity which wanes over time returning them to the susceptible state.

In the model, IPTp is administered to all pregnant women unless they are receiving antimalaria treatment, in which case they will receive IPTp upon recovery. The model captures four stages of zero, one, two and three dose(s) of IPTp administered to pregnant women based on WHO recommendations of at least three doses of IPTp [2]. A proportion of pregnant women with zero doses of IPTp receive the first dose of IPTp at the beginning of the fourth month of pregnancy and transition to the 1-dose protected states (*P*_1,*R*_ and *P*_1,*S*_). They remain in this state for a maximum period of 42 days (4 to 6 weeks) [28]. After one month, a proportion of pregnant women (in the 1-dose states) may receive the second dose of IPTp and enter the 2-dose protected states. The other proportion of pregnant women who did not receive the second dose of IPTp will lose IPTp protection once the protected period ends and become vulnerable to infection again. Finally, a proportion of pregnant women (in the 2-dose states) may receive the third dose of IPTp and transition to the 3-dose protected states.

A diagrammatic representation of the mathematical model of malaria incorporating 0 doses, 1 dose, 2 doses, and 3 doses of IPTp is shown in Fig 1. The model state variables, parameters, and the corresponding equations are fully described and outlined in Table S1, Table S2 and Equations (1) - (38), respectively, in the supplementary file.

**Fig 1.**
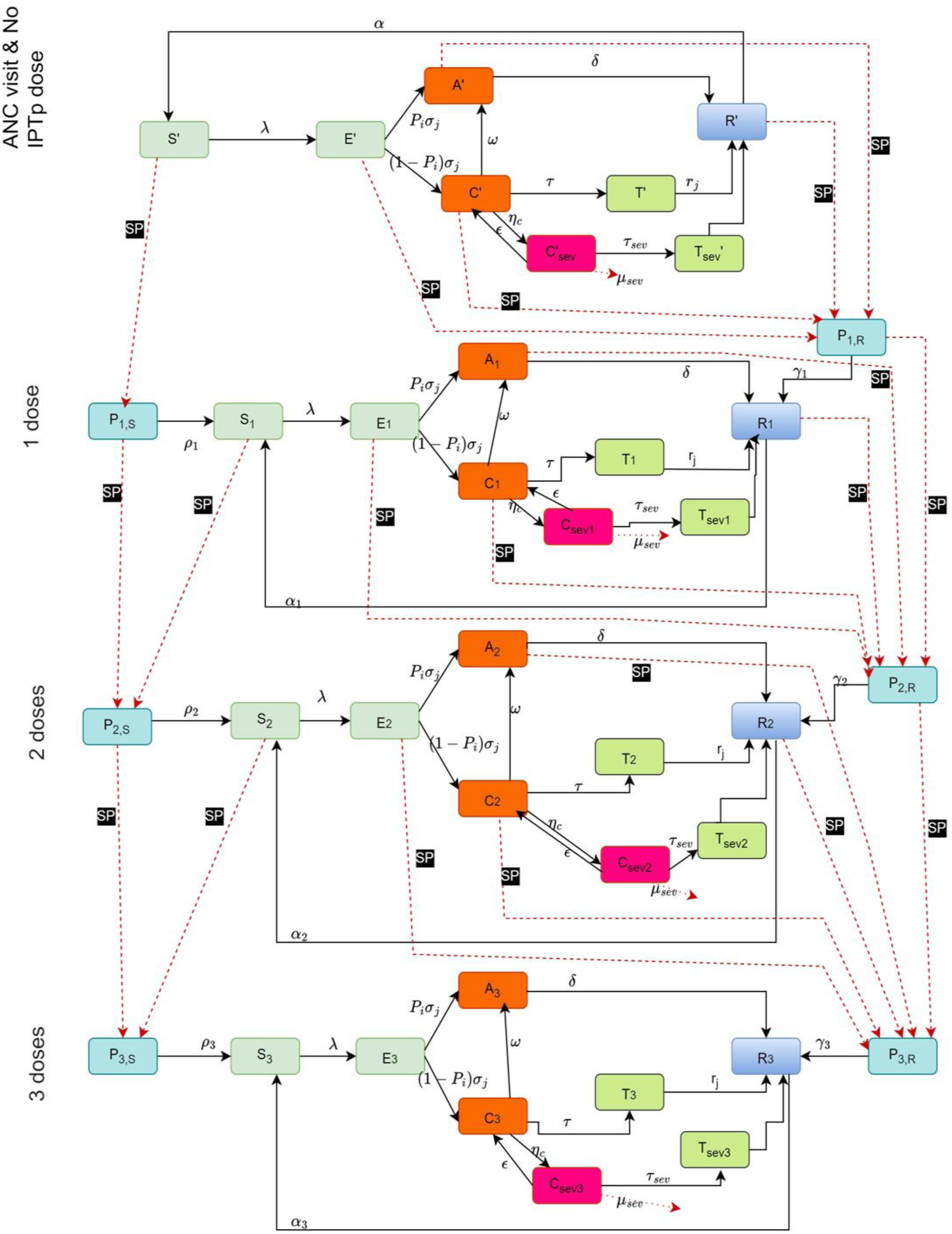
Flow diagram for malaria model including implementation of 0, 1, 2 and 3 doses of IPTp with sulfadoxine-pyremethamine (IPTp-SP) as prophylaxis for a cohort of pregnant women. IPTp-SP administration flows are represented by dashed lines with the SP symbol.

In the mathematical model in Fig 1, the following assumptions are made:

- The population in the model is a cohort of pregnant women who have access to ANC services.
- All pregnant women, irrespective of disease status, have access to IPTp during ANC visits, with a proportion of them receiving three doses of IPTp.
- The cohort being modeled consists of women who all began their pregnancies in the same month. We will explore scenarios of different cohorts starting their pregnancies at different months of the year. There is no mid-way recruitment, meaning no new participants are added after the initial start. The model tracks the days, weeks, and months of pregnancy from day one for all participants, without interaction with those who start their pregnancies at different times.
- Pregnant women are exposed to the same transmission reservoir as other individuals since they live in a larger population. Thus, our cohort of pregnant women contract malaria at a constant force of infection (FOI) of the general population within which they live in, which is described in the supplementary file. This implies that any infectious mosquitoes that are present in the larger community will also have equal interaction with pregnant women who live there.
- Each woman who has received a dose of IPTp has a risk of becoming susceptible again due to the imperfect efficacy of IPTp. Other prevention methods such as ITNs can be used to prevent infection among these susceptible sub-groups.
- After ten months, when the women are no longer pregnant, they are removed from the active population regardless of doses received.

### Data

Publicly available data was used to parameterize the mathematical model. Total ITN distribution data for Zimbabwe was obtained from the World Malaria Report annexes for a period of ten years (2013 – 2023). The data was used to estimate ITN coverage and incorporate the effects of ITNs into FOI. National rainfall daily data for Zimbabwe for the period 2013 to 2023 was downloaded from the Climate Hazards Group InfraRed Precipitation with Station data (CHIRPS) website. We used the rainfall data to account for the effects of seasonality. Rainfall patterns affect the seasonal phases of transmission as well as mosquito breeding cycles [25, 29]. The mathematical model is illustrated by Equations (1) - (38) in the supplementary file and was programmed using R version 4.2.3 software.

### Intervention scenarios

The baseline scenario for the IPTp model represents the average coverage levels of one, two and three doses of IPTp in Southeast Africa, which we obtained from the World Malaria Report [11]. These values were applied to estimate the baseline IPTp dose coverages in the model. Other hypothetical scenarios were created, with a focus of reducing the burden of malaria in pregnancy and the associated birth outcomes of LBWs. We explored the following scenarios for IPTp and ITN use interventions as shown in Table 1.

**Table 1.** Intervention scenarios showing the coverages of IPTp 1, 2 and 3 doses and ITN use across baseline and hypothetical alternatives focusing on reducing the burden of MiP and LBWs.

| Scenario | Description |
| --- | --- |
| Baseline | IPTp1 64%, IPTp2 54%, IPTp3 42% |
| High IPTp low ITN | Increasing IPTp1, IPTp2 and IPTp3 to 90%, wider ITN use during pregnancy 50% |
| High IPTp medium ITN | Increasing IPTp1, IPTp2 and IPTp3 to 90%, wider ITN use during pregnancy 70% |
| High IPTp high ITN | Increasing IPTp1, IPTp2 and IPTp3 to 90%, wider ITN use during pregnancy 90% |
| Low IPTp medium ITN | Increasing IPTp1 to 90%, while decreasing IPTp2 to 50%, IPTp3 to 40%, wider ITN use during pregnancy 70% |

### Pregnancy cohorts’ scenarios

For the pregnant women, the timing of the peak in rainfall is very important because higher malaria transmission periods bring a greater risk of infection to them [30, 31]. We introduced a parameter in the model to shift the start of pregnancy in calendar time while keeping the seasonal transmission cycle unchanged. The model was parameterised using rainfall data for Zimbabwe, where rainfall and consequently malaria transmission usually peak in January. In these scenarios, cohorts of pregnant women begin pregnancy at different times of the year, resulting in different stages of gestation overlapping with the seasonal peak in transmission. The scenarios are fully described in Table 2. Each cohort was scheduled to receive three doses of IPTp, with all doses administered by the end of the sixth month of pregnancy.

**Table 2.** Pregnancy cohorts defined by the month in which pregnancy begins and the corresponding gestational stage at the January seasonal malaria transmission peak occurring during or after pregnancy has started. following January. For pregnancies which started between February and December, the January transmission peak refers to the following January.

| Month pregnancy begins (scenario) | Gestational age and trimester of pregnancy at the January transmission peak |
| --- | --- |
| January | Pregnancy has just begun |
| February | Pregnancy completed before subsequent transmission peak |
| March | Ten months, third trimester (close to delivery) |
| April | Nine months, third trimester |
| May | Eight months, third trimester |
| June | Seven months, third trimester |
| July | Six months, second trimester |
| August | Five months, second trimester |
| September | Four months, second trimester |
| October | Three months, first trimester |
| November | Two months, first trimester |
| December | One month, first trimester |

### Model checks

We conducted sensitivity analysis to assess whether the model behaved as expected by varying selected parameters and observing changes in the total cumulative incidence. Further details of the sensitivity analysis are provided in the Supplementary Material (Fig S3, Table S4 and Table S5). For parameters with uncertainty, probability distributions were assigned based on values reported in the literature (Table S3), and parameter intervals were defined accordingly. Random values within these intervals were sampled from their respective distributions, and 100 simulations were run in R for all scenarios. The results were then summarised by calculating the median, 5th and 95th percentiles of the uncertainty interval.

## Results

We considered moderate and high malaria transmission intensities discussed earlier in the study settings, to compare the dynamics of malaria infection during pregnancy and to estimate the effects of IPTp and pregnancy timing on cases of MiP and LBWs. Our analysis was divided into three parts. First, we presented the results of the analysis on how the intervention scenarios affect clinical malaria cases and LBW outcomes from malaria, among the August cohort of pregnant women (whose pregnancy started in August). Next, we analysed cohorts with varying pregnancy start times to examine how the overlap with peak incidence periods affects clinical cases and LBW risk. Finally, we analysed the effects of intervention scenarios across the different pregnancy cohorts.

### Effects of interventions

Fig 2 shows the estimated monthly number of clinical cases in moderate and high transmission settings for a cohort whose pregnancy starts in August. In Fig 2, the baseline scenario with colour red has the highest monthly number of clinical cases throughout the pregnancy period as compared to the other scenarios in both moderate and high transmission. The high IPTp high ITN scenario has the lowest number of monthly clinical cases. The plot also shows that, for the same high IPTp coverages, the lower the wider ITN use during pregnancy is associated with higher clinical cases. This provides evidence on the effectiveness of ITN use in preventing malaria infections. The low IPTp medium ITN scenario shows cases increasing from month 4 to 6. This is likely due to the low coverages of IPTp resulting in fewer pregnant women becoming protected during this period.

**Fig 2.**
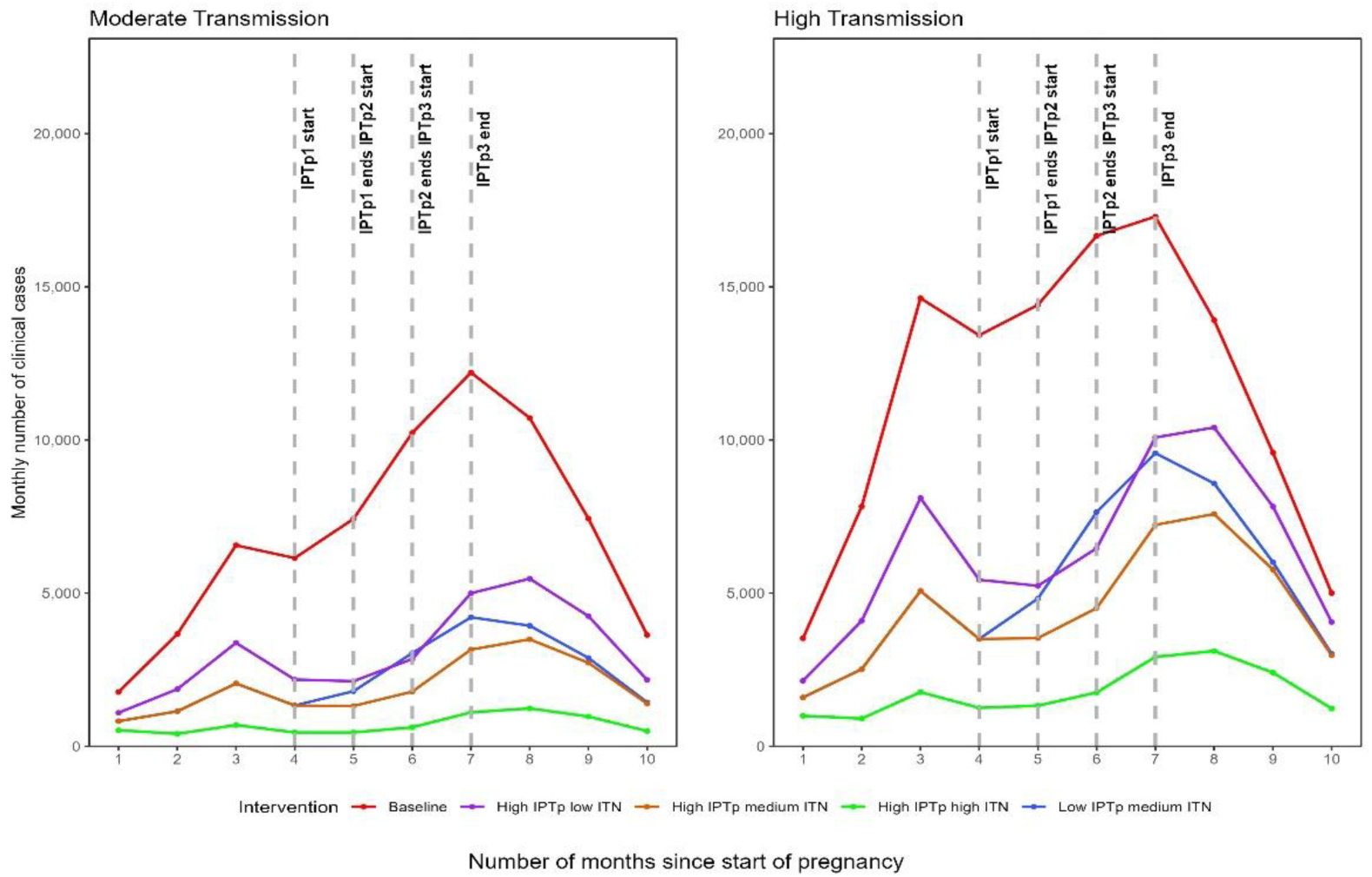
Estimates of the monthly number of clinical malaria cases in moderate and high transmission settings under different intervention scenarios for baseline pregnancies that started in August. The monthly points display average number for the clinical cases.

Furthermore, among scenarios with the same medium ITN but different IPTp coverages, higher IPTp coverage was associated with fewer total clinical cases. For example, under moderate transmission, high IPTp coverage reduced total clinical cases to 18,989, compared with low IPTp coverage; under high transmission, the corresponding reduction was to 43,761. The full comparison of total clinical cases for the 10-month simulation period across all intervention scenarios relative to the baseline for the August cohort is shown in the supplementary file (Table S5).

The calculation of total number of LBW cases resulting from the different intervention scenarios was done using an approach described in the supplementary file (Fig S5). Moreover, using our mathematical model, we estimated the percentage of LBWs averted by different scenarios relative to the baseline scenario. Fig 3 shows that high IPTp high ITN, which has the highest coverages of IPTp at 90% and high ITN use during pregnancy, averts of 73% and 71% of LBWs in moderate and high transmission settings, respectively. Our findings suggest that low IPTp medium ITN scenario, in which only IPTp1 coverage is high at 90%, while IPTp2 and IPTp3 coverage remain low and ITN use during pregnancy is moderate, prevents 44% and 37% of neonates with LBWs in moderate and high transmission settings. These percentages of LBWs averted are lower compared to those observed where coverage of all three IPTp doses remains high.

**Fig 3.**
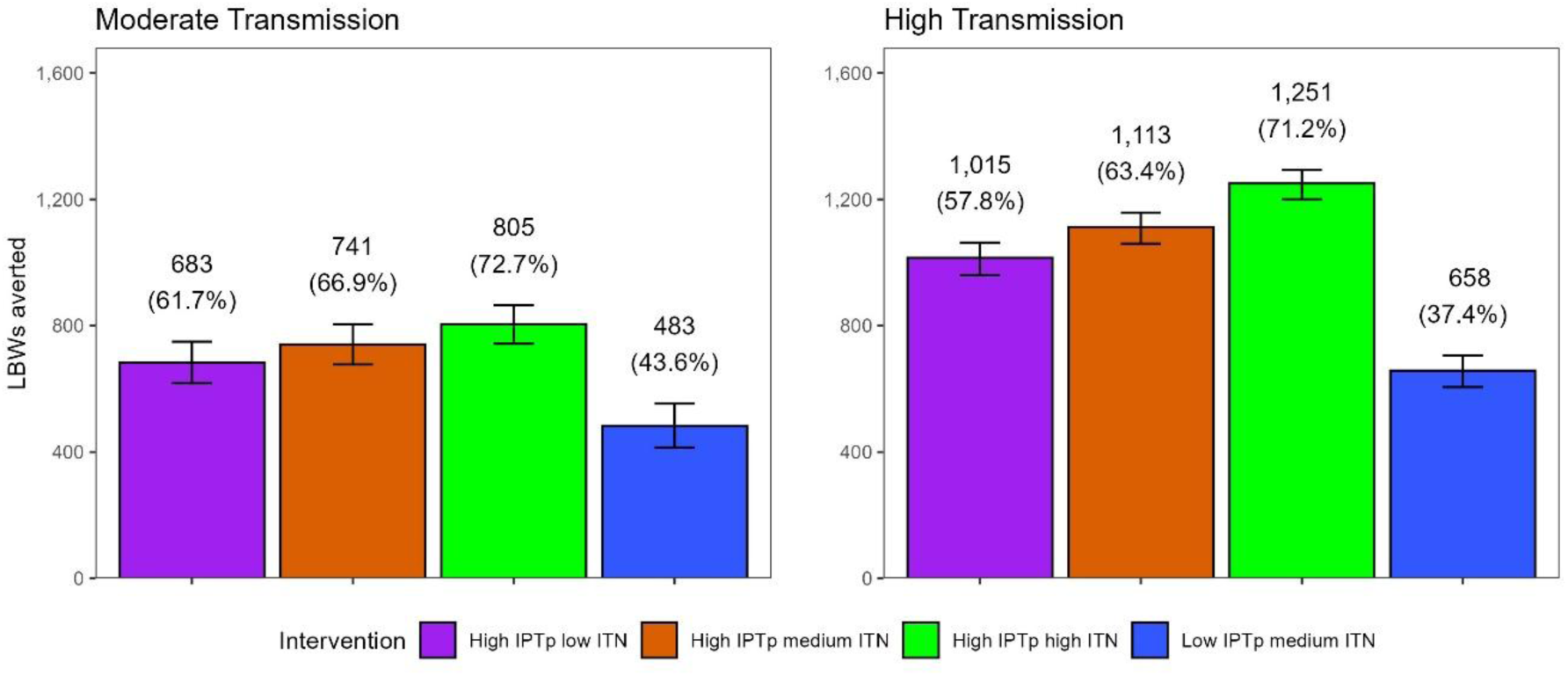
Predicted number and percentages of LBWs averted by different combinations of interventions in moderate and high transmission setting at the end of the ten-month pregnancy period for pregnancies that started in August. Intervention scenarios are defined in Table 1. Scenarios were compared against the baseline to assess LBW cases averted. Bars show the average number of LBW cases averted and their respective 90% uncertainty interval derived from parameter variability.

### Effects of the timing of pregnancy and seasonality peak

We created scenarios of cohorts with different pregnancy start times to assess the effect of pregnancy timing in relation to seasonal transmission and these are described in detail in Table 2. Our model was parameterised with data from Zimbabwe which has peak rainfall in January. Fig 4 shows the monthly numbers and total numbers of clinical cases among twelve different cohorts of pregnant women in moderate and high transmission settings. The plot illustrates how the number of clinical cases varies depending on when the pregnancy started. Fig 4A shows that monthly number of cases are generally lowest in the months 4 to 6 across all cohorts in both transmission settings. These results correspond to the period when the pregnant women would be receiving IPTp doses in our model and in protected states. However, since only proportions of pregnant women would be receiving IPTp, some pregnant women can still become infected and have clinical cases in the months when IPTp is being delivered.

**Fig 4.**
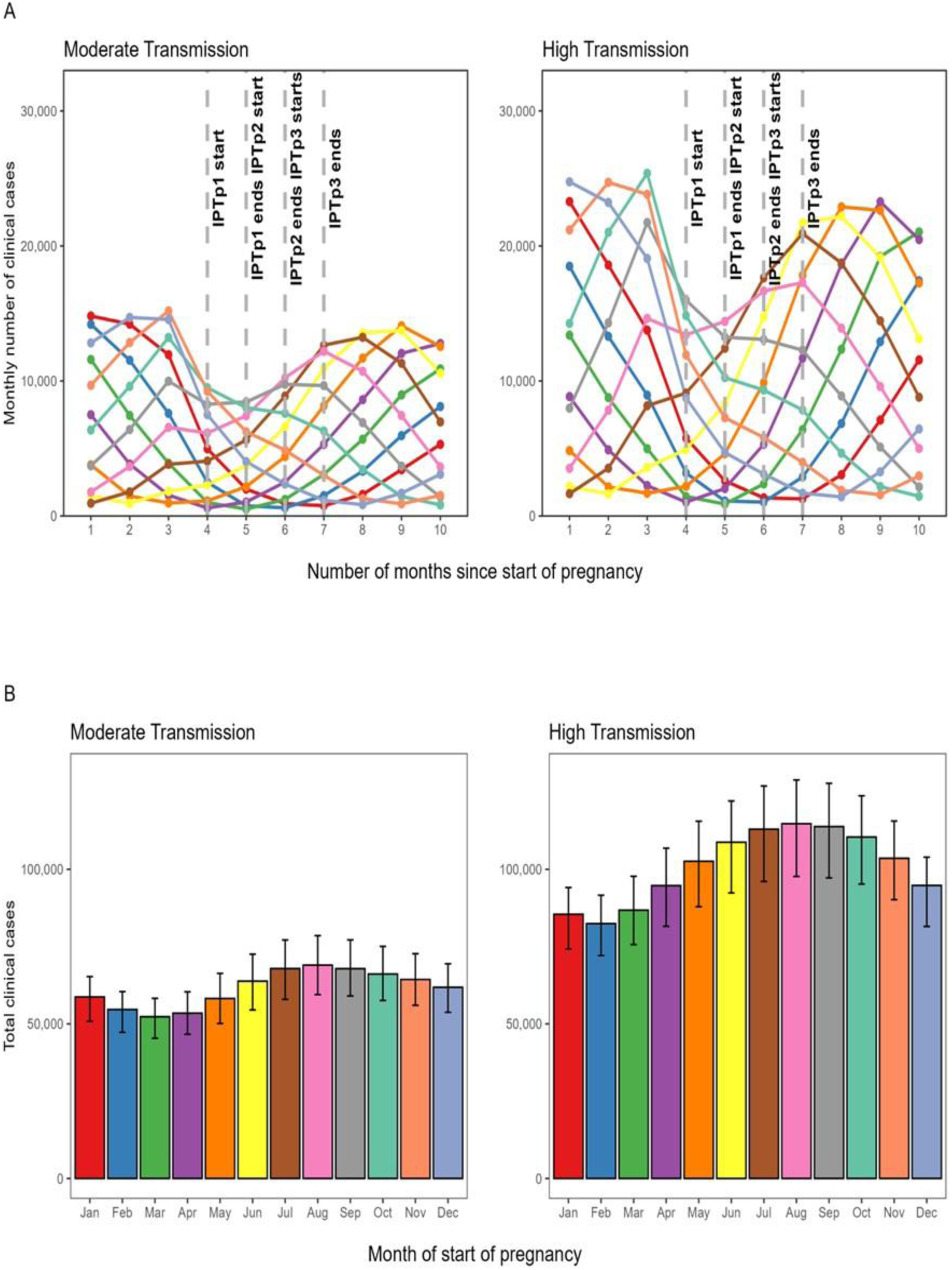
Estimates of monthly number of clinical cases and the total number of clinical cases for the different pregnancy cohorts described in Table 2 at the end of the 10-month simulation period. The plot shows results for the baseline intervention scenario only (IPTp1 64%, IPTp2 54%, IPTp3 42%). A. Comparison of monthly clinical cases in high and moderate transmission settings for different scenarios of pregnancy start times. B. Comparison of the total number of clinical cases for different pregnancy start times for the cohorts. Bars show the average number of clinical cases and their respective 90% uncertainty interval derived from parameter variability.

Conversely, Fig 4B shows that the lowest total number of clinical cases was recorded in the March cohort and February cohort in moderate and high transmission settings respectively. These pregnancies were nearing delivery or had ended in January when malaria incidence is high. However, the highest total number of clinical cases are recorded in the August cohort in both moderate and high transmission, which overlaps with the January period of high malaria incidence at the end of their second trimester. This is because towards the end of the second and during the third trimesters in our simulation, most of the pregnancies have lost the protection from IPTp. Our findings imply that the August cohort of women need to start to take the doses very early in their second trimester until delivery otherwise they would be at higher risk of infection.

The estimated number of neonates with LBW from different cohorts of pregnant women is shown in Fig 5. Our results highlight that at the baseline intervention scenario, under both moderate and high transmission, the lowest number of neonates with LBW was recorded for the December cohorts. These cohorts have pregnancies which overlap with the January peak of malaria infections during the first trimester. On the other hand, in moderate transmission, the highest number of neonates with LBW was recorded for the June cohort, which overlapped with the peak of malaria infections during the third trimester. These pregnancies experienced an increased number of infections in the third trimester when they were unprotected. On the contrary, the highest number of neonates with LBW was recorded for the May cohort in high transmission. This cohort had pregnancies which overlapped with the peak phase of malaria incidence towards the end of the third trimester. The high numbers of neonates with LBW recorded in the third trimester could be because, at that stage and at the baseline intervention scenario, our cohorts have lost all protection from IPTp since they were scheduled to receive only three doses. Thus, the May and June cohorts require more than the standard three doses of IPTp to sufficiently reduce the chances of becoming infected and subsequently having a LBW baby.

**Fig 5.**
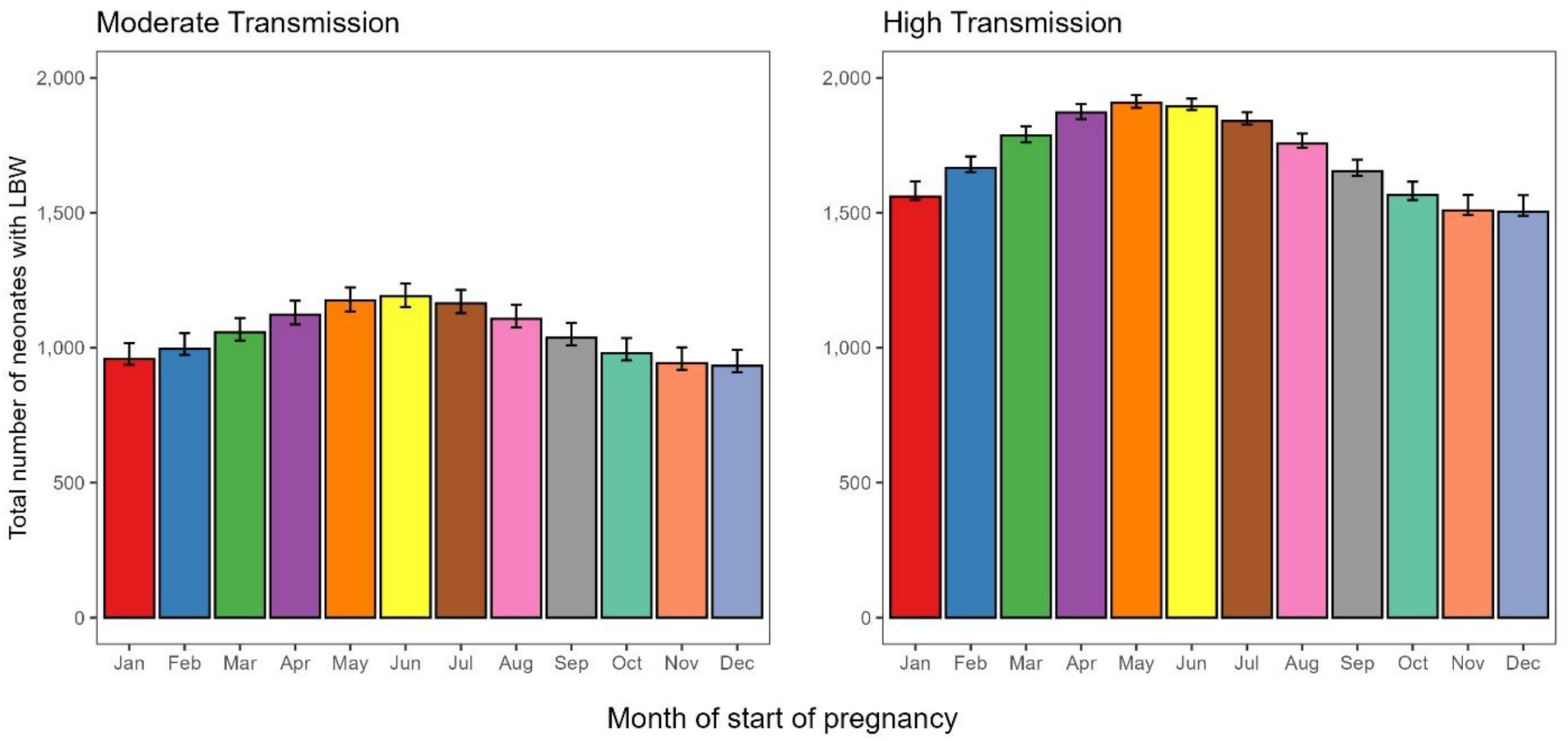
Estimated number of neonates with LBW for different pregnancy start times cohorts in moderate and high transmission settings. The plot shows results for the baseline intervention scenario (IPTp1 64%, IPTp2 54%, IPTp3 42%). Bars show the average number of neonates with LBW and their respective 90% uncertainty interval derived from parameter variability.

### Effects of interventions in different pregnancy cohorts

Fig 6 shows that the high IPTp high ITN scenario, where increasing IPTp coverage for all three doses to 90% and high wider ITN use during pregnancy leads to very low numbers of clinical cases for all pregnancy cohorts in both moderate and high transmission settings. This clearly provides strong evidence of the effectiveness of both increased IPTp and ITNs in reducing clinical cases of malaria. The plot shows that High IPTp low ITN scenario, despite higher IPTp coverages, results in a higher number of clinical cases compared with the Low IPTp medium ITN scenario (IPTp1 = 90%, IPTp2 = 50%, IPTp3 = 40%, ITN = 70%). This suggests that increased ITN use during pregnancy plays a critical role in reducing clinical malaria cases. The results further show that increasing IPTp coverage contributes to reducing clinical cases. This is shown when comparing the scenarios with medium wider ITN use during pregnancy but different IPTp coverages. The scenario with higher IPTp coverage results in fewer clinical cases than the scenario with low IPTp coverage. Nonetheless, the overall pattern illustrates the effectiveness of increasing IPTp coverage in reducing clinical cases.

**Fig 6.**
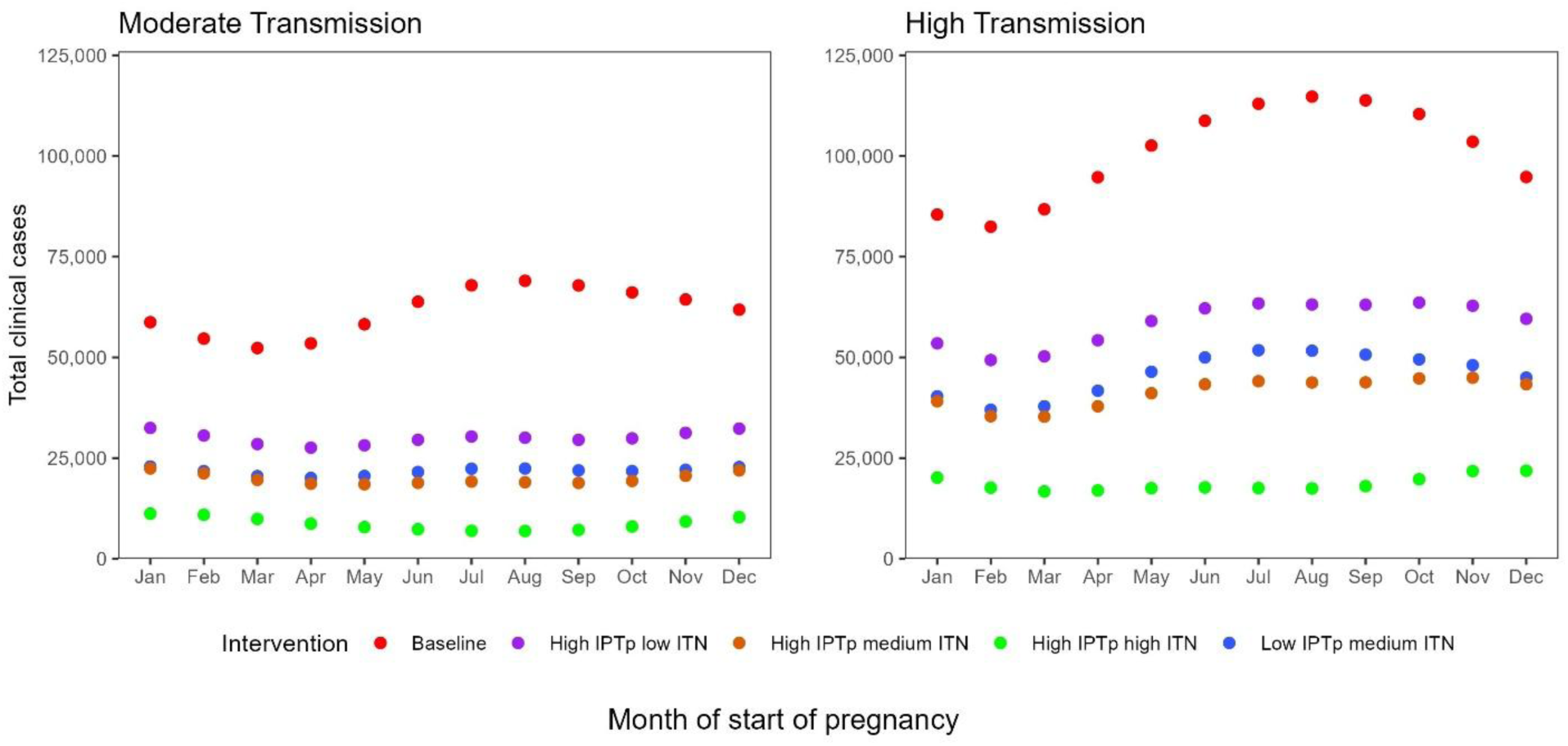
Estimated total number of clinical malaria cases for different pregnancy start times across different intervention scenarios under moderate and high transmission settings. The points represent the average number of total clinical cases. The intervention scenarios are fully described in Table 1.

Our results indicate that, high IPTp and high ITN use during pregnancy averts an average of 86% to 81% of clinical cases compared with the baseline for all the pregnancy cohorts, in moderate and high transmission respectively. In contrast, the scenario with low ITN use during pregnancy averted few clinical cases when compared to the other scenarios in both transmission settings and across all pregnancy cohorts (averages of 51% in moderate and 41% in high transmission settings). More details on clinical cases averted are available in Fig S5 in the supplementary file.

Fig 7 shows that the scenario with both high IPTp coverages and high ITN use during pregnancy has the lowest numbers of LBWs compared to other scenarios varying levels of ITN use during pregnancy in both moderate and high transmission settings. This provides strong evidence that high IPTp coverage is very effective in preventing LBWs when implemented alongside high ITN use during pregnancy. The plots illustrate that low IPTp medium ITN scenario results in higher numbers of LBWs compared to the high IPTp low ITN scenario. This pattern differs from that observed for the same scenarios when considering clinical cases. This provides evidence of the effectiveness of IPTp in combating LBWs. Our results imply that increasing the coverage of IPTp1 is very impactful, and early administration in the second trimester is important due to the elevated risk of infection sequestering within the placenta. However, there is a need to sustain IPTp2 and IPTp3 through improved ANC attendance and follow ups. The percentages of LBW cases averted by the intervention scenarios compared to the baseline for all the different pregnancy cohorts are illustrated in Fig S6 in the supplementary file.

**Fig 7.**
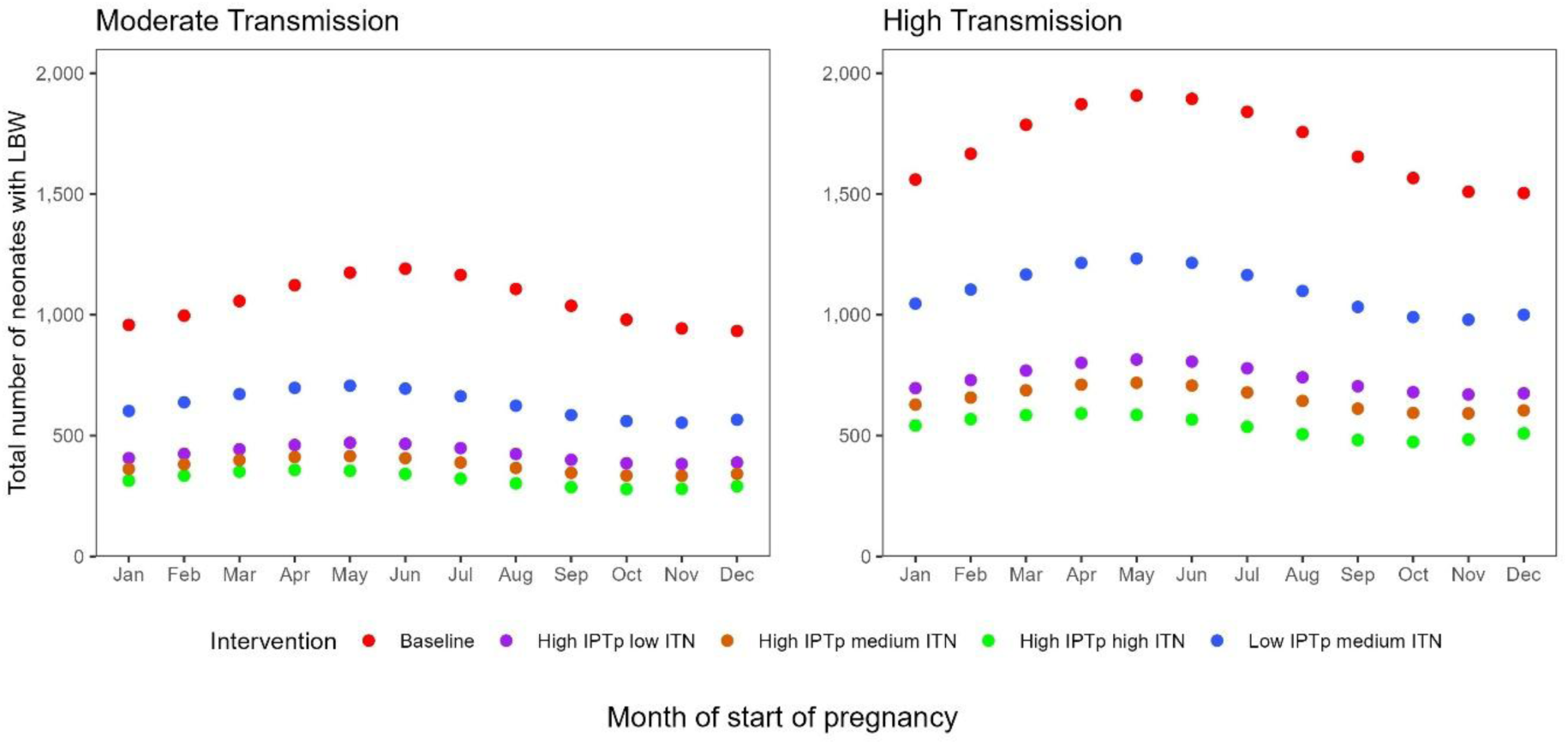
Estimated number of neonates with LBW at the end of the ten-month simulation period for pregnancies with different start times across different intervention scenarios defined in Table 1 under moderate and high transmission. The points represent the average number of total number of neonates with LBW. The intervention scenarios are fully described in Table 1.

## Discussion

Malaria during pregnancy remains a major contributor to poor birth outcomes and a leading cause of maternal and newborn mortality in Sub-Saharan Africa. This calls for investments in malaria control programs and ANC health systems that can quicken the eradication of the infection. In this study, we formulated a mathematical model of MiP, which included IPTp, ITNs, and seasonality, and used this model to simulate the dynamics of malaria transmission in moderate and high transmission environments. Publicly available data for Zimbabwe was used to parameterise the model. The simulation findings demonstrate how well IPTp and ITNs work to lower the burden of MiP and LBWs. To reduce maternal and newborn problems associated with malaria, our model results emphasize the necessity of improved ITN use during pregnancy and high IPTp coverage.

Our results show that increasing the coverage of the first dose of IPTp to 90% (exceeding first ANC attendance rates [3, 4, 11]), while maintaining lower coverage for subsequent doses, prevents an average of 40% and 35% of LBW cases across all pregnancy cohorts compared with the baseline intervention scenario, in moderate and high transmission settings, respectively. Our finding shows that high IPTp1 coverage is the most impactful call for improvement in the health system to address absconding of IPTp dose uptake. By increasing IPTp uptake to levels above first ANC attendance, the incidence of LBW, due to malaria, could be significantly decreased [6, 14]. The incremental benefits of expanding IPTp-SP coverage are nevertheless substantial even in regions with greater levels of SP resistance, even though geographic differences in SP resistance somewhat reduce the effectiveness of IPTp-SP [4, 6, 14, 25, 29, 32, 33]. However, the coverage of IPTp remains very low in many sub-Saharan African countries [4, 34, 35].

In our simulations, the greatest decrease in LBW and clinical cases, for all pregnancy cohorts, was predicted to be when high coverages of IPTp and ITNs were used in tandem (compared to when only IPTp coverages are increased). Previous research supports this dual approach by showing that ITNs provide crucial environmental protection by lowering exposure to mosquito bites, while IPTp efficiently addresses the biology side of treating malaria during pregnancy [14]. When both strategies are used together, the risk of neonates with LBW and malaria infection is reduced more efficiently than when one is used alone [14, 36]. However, analysis of the World Malaria Report annexes for the moderate to high transmission countries implementing IPTp shows very low percentages of pregnant women using ITNs and/or who took at least 3 doses of IPTp [3, 4, 11, 12]. Furthermore, our results demonstrate that scenarios with high IPTp coverage and medium ITN use prevented the highest number of clinical cases and LBWs compared to scenarios with lower IPTp coverage at similar levels of ITN use. These findings highlight the important role of increasing IPTp coverage in substantially reducing the burden of MiP and lowering clinical malaria cases during pregnancy, even when ITN use remains at moderate levels. These findings agree with previous studies, which found that IPTp had high LBWs prevention benefits, whilst its efficacy in reducing malaria infections has been slightly reduced due to resistance [33, 37]. On the contrary, our results highlight that low IPTp coverages and medium ITN use averted higher numbers of clinical cases when compared to the high IPTp low ITN use. The results demonstrate the importance of scaling up ITN distribution and use during pregnancy. These findings agree with those in literature which found that ITN use during pregnancy had a beneficial impact on MiP and its outcomes [38, 39].

In addition, our study shows that variation in pregnancy timing and peak period of malaria transmission had a significant impact on the total disease burden. This observation aligns with previous research, which showed that the number of infections recorded varies according to when the pregnancy started [9, 15]. We found that pregnancies started in August faced the increased risk of clinical infections at the baseline intervention scenario because they overlapped with the January seasonality peak during the second trimester. On the contrary, pregnancies which started in May and June faced the highest burden of LBWs for the baseline intervention scenario because they overlapped with the January seasonality peak during the third trimester. These findings imply that early initiation of IPTp in the second trimester is recommended for the May, June and August pregnancies. At the baseline scenario (with decreasing IPTp coverages), these pregnancies would need more than three doses of IPTp to be fully protected. This finding ties well with previous research and WHO recommendations that pregnancies should be protected from the beginning of the second trimester until delivery to prevent infections and improve birth outcomes [2]. However, our simulations demonstrate that when there are high IPTp coverages and high ITN use during pregnancy the burden from MiP is significantly reduced across all pregnancy cohorts. This finding suggests that timely administration of IPTp doses in higher coverages, combined with consistent ITN use, can provide strong protection throughout pregnancy, regardless of when the pregnancy started.

This study has its strengths, but also limitations that need to be considered when interpreting the findings. The uniqueness of this study is that it examines the overlap of seasonality phase and the gestational ages for different cohorts of pregnant women in moderate and high transmission settings. The findings of this study have clear implications for policymakers and can be used for effective planning and rolling out of the interventions for MiP. The research adds to existing literature that supports the idea that scaling up of IPTp has a significant effect on the numbers of neonates with LBWs and thus provides long-term benefits. Our simulations can be used to provide evidence on the number of IPTp doses which can be distributed to a given population of pregnant women at stated coverage levels.

At the same time, one limitation of our study was that we had limited data on the number of pregnant women who received IPTp broken down per dose, on those who had malaria infections during their pregnancy and on malaria-related LBWs in these transmission areas. Availability to this malaria data would have enabled the validation of the mathematical model through comparison of the simulated clinical infections and LBWs to the observed patterns. Thus, this could have led to overestimation or underestimation of clinical cases and LBWs. Future research should reconfirm these findings by following the pregnant women throughout their pregnancy and conducting data collection. Hence, caution should be taken in generalizing the findings to all moderate and high transmission areas in Southeast Africa as some results may not apply to some countries.

Despite these limitations, this study highlighted useful insights on increasing IPTp coverage and getting the IPTp doses until delivery. Future work could explore the effects of resistance on IPTp effectiveness over time and the outcomes of MiP. We plan to evaluate the use of alternative IPTp delivery pathways to improve uptake in the future.

## Conclusions

This study highlights the benefit of scaling up IPTp uptake and ITN use among pregnant women. The results show that increasing the coverage of IPTp doses and high ITN use prevents a higher number of neonates with LBWs, which, in turn, reduces the chances of malaria-related retarded growth. Our study also demonstrated that exposure to malaria and the risk of LBWs varies depending on the time of the year the pregnancy started and its overlap with the rainy season. Pregnancies beginning in August for both moderate and high transmission experienced the highest burden of total clinical cases. The highest burden of LBW outcomes was observed among pregnancies whose third trimester coincided with the January seasonal transmission peak. Results of the study are key because they underscore the cruciality of improving provision, administration and promoting the use of malaria preventive measures among pregnant women until delivery.

## Data Availability

The data used in this study are open access and freely available from the respective sources.

## Notes

### Competing Interest Statement

The authors have declared no competing interest.

### Author Declarations

The study was cleared for ethical considerations by the University of Cape Town (UCT) research ethics committee (HREC REF 179/2024). Furthermore, the study`s protocol was reviewed and approved by the full board of the Medical Research Council of Zimbabwe (approval number: MRCZ/A/3363).

### Summary of Updates

The version of the manuscript has been updated to include the supplementary file which was previously not attached.

