## Supplemental File for "Estimating the impact of different Intermittent Preventive Treatment in Pregnancy delivery strategies on low birth weight outcomes under moderate and high malaria transmission settings: A modelling study"

**Supplementary file**

**Mathematical model formulation and description**

The standard process of how pregnant women living in the high malaria burden areas of Zimbabwe receive IPTp at ANC is illustrated in Fig S1.

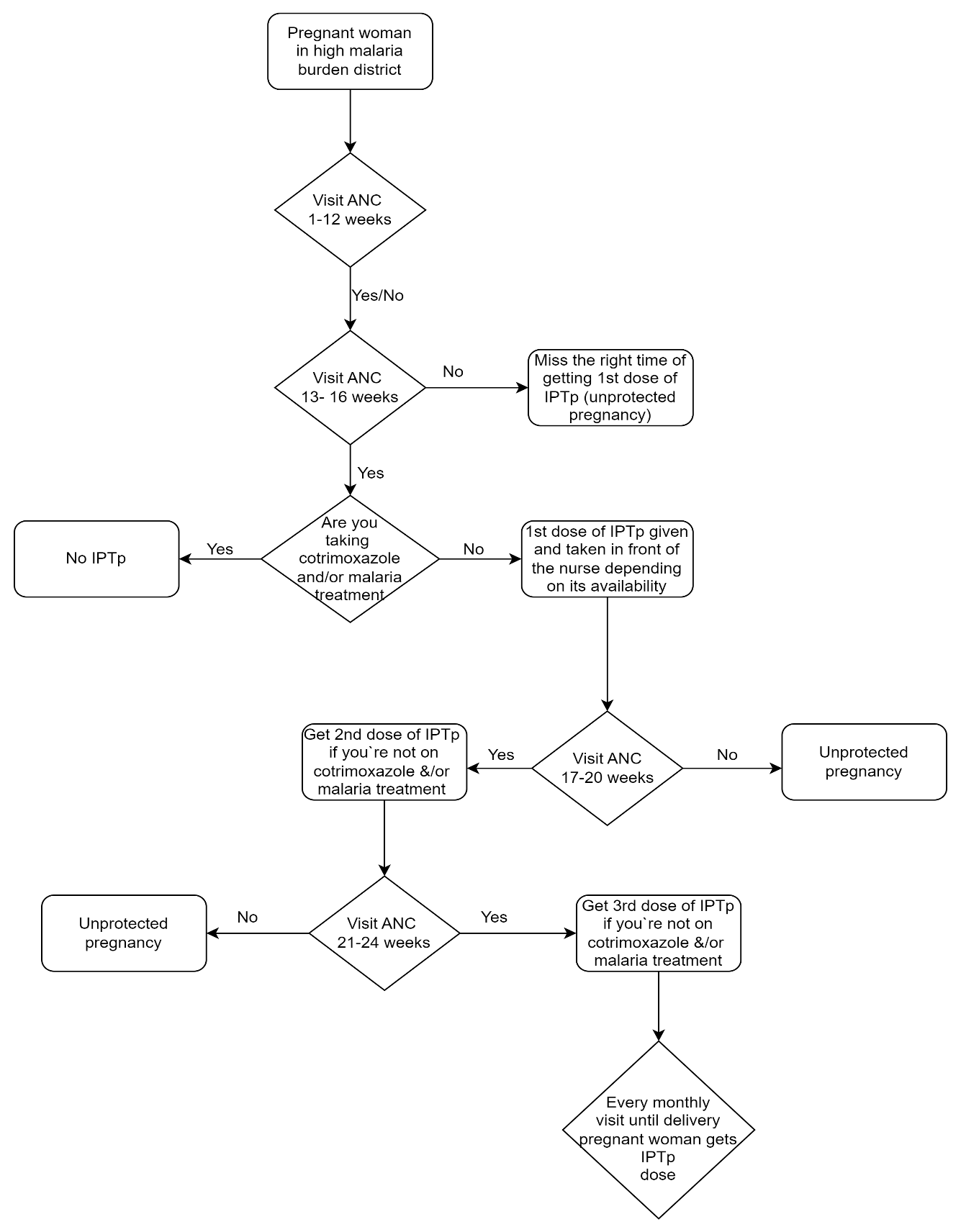

**Fig S1. Flow chart for the IPTp process in Zimbabwe.**

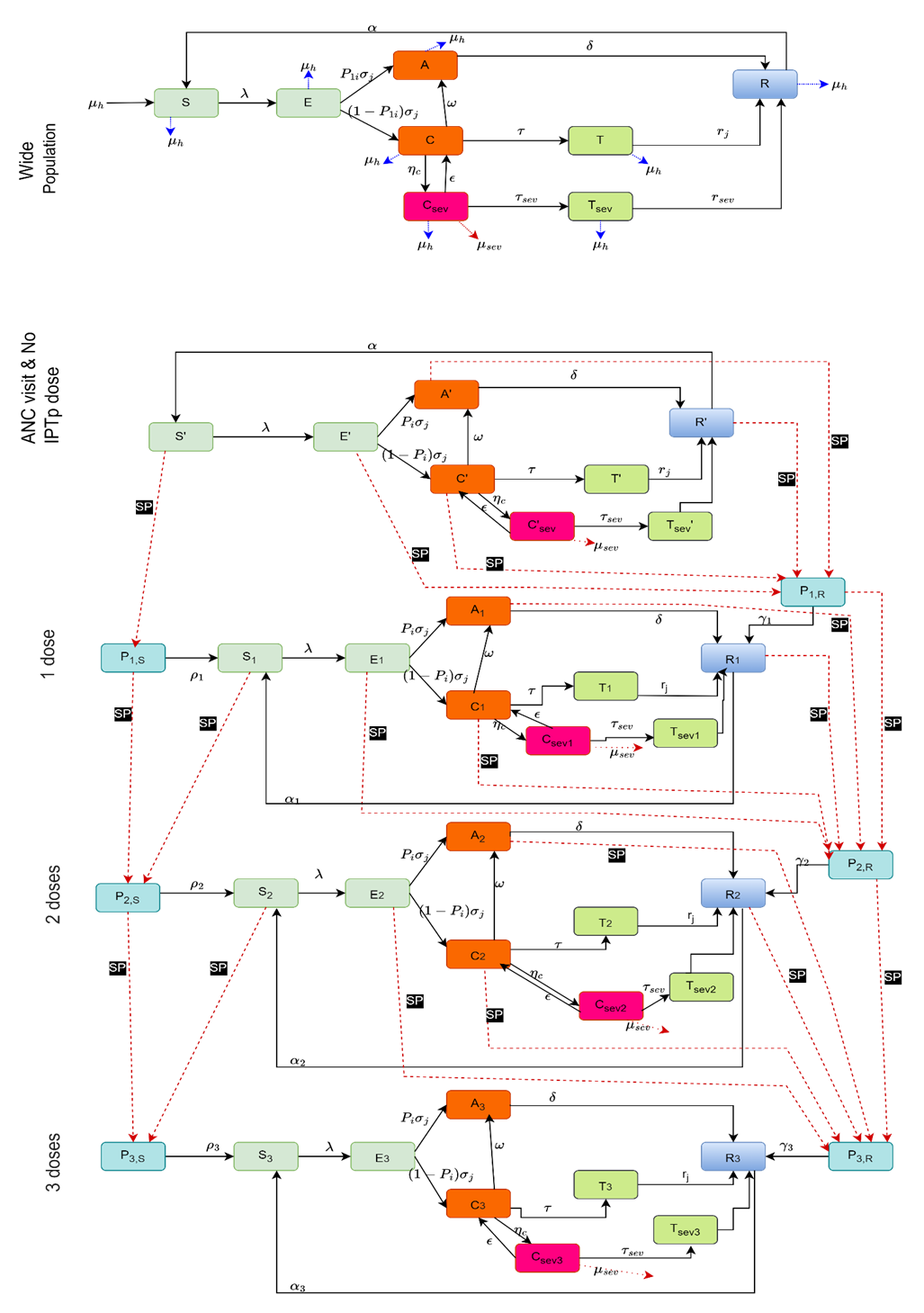

**Fig S2. Malaria transmission model showing wide population model and the cohort model of malaria in pregnancy with IPTp intervention.** The state variables and parameters are described in Table S1 and Table S2.

The model in Fig S2 was developed to reflect the malaria dynamics among the wide population in which the pregnant women live in and among the cohort under study. People are recruited into the wide population model by birth. The population in the susceptible class are uninfected and they can become infected at a rate of λ. Recovered individuals lose their immunity to become susceptible again at a rate of α. When exposed to malaria (after an infectious mosquito bite), individuals go through the incubation period before becoming infectious. Infectious individuals can either be asymptomatic, with clinical symptoms or severely ill. The population with asymptomatic malaria has the disease but they do not have symptoms, and they recover naturally at a rate of δ. Infectious individuals with clinical symptoms can either seek treatment, progress to severe malaria or lose the clinical symptoms without treatment and become asymptomatic. A proportion of severely ill individuals can be hospitalized while the other proportion can die due to complications of severe disease or lose severity and become clinical. Treated individuals lose their infectiousness and enter the recovered class. Individuals in the wide population die naturally at a rate of $\mu_{h}$.

A simulated cohort of pregnant women is generated from the wide population model and goes through the same transmission dynamics that the individuals in the wide population go through. Those in susceptible or uninfected compartments get infected at the same rate λ as those in the wide population. The dynamics of malaria among our simulated cohort and the administration of IPTp has been described in the main file.

The state variables for the model of IPTp are fully described in Table S1.

**Table S1. State variables for the model.** $i$ represents 1, 2 and 3.

| State variable | Symbol |
| --- | --- |
| S | Susceptible population |
| E | Exposed population |
| A | Asymptomatic population |
| C | Clinical population |
| $\boldsymbol{C}_{\boldsymbol{sev}}$ | Severely ill population |
| $\boldsymbol{T}_{\boldsymbol{sev}}$ | Treated population who had severe malaria |
| T | Treated population who had clinical malaria |
| R | Recovered population |
| S` | Susceptible pregnant women with zero doses of IPTp |
| E` | Exposed pregnant women with zero doses of IPTp |
| A` | Asymptomatic pregnant women with zero doses of IPTp |
| C` | Clinical pregnant women with zero doses of IPTp |
| $\boldsymbol{C`}_{\boldsymbol{sev}}$ | Severely ill pregnant women with zero doses of IPTp |
| $\boldsymbol{T}_{\boldsymbol{sev}}\boldsymbol{`}$ | Treated pregnant women who had severe malaria with zero doses of IPTp |
| T` | Treated pregnant women who had clinical malaria with zero doses of IPTp |
| R` | Recovered pregnant women with zero doses of IPTp |
| $\boldsymbol{S}_{\boldsymbol{i}}$ | Susceptible pregnant women with $i$ doses of IPTp |
| $\boldsymbol{E}_{\boldsymbol{i}}$ | Exposed pregnant women with $i$ doses of IPTp |
| $\boldsymbol{A}_{\boldsymbol{i}}$ | Asymptomatic pregnant women with $i$ doses of IPTp |
| $\boldsymbol{C}_{\boldsymbol{i}}$ | Clinical pregnant women with $i$ doses of IPTp |
| $\boldsymbol{C}_{\boldsymbol{sevi}}$ | Severely ill pregnant women with $i$ doses of IPTp |
| $\boldsymbol{T}_{\boldsymbol{sevi}}$ | Treated pregnant women who had severe malaria with $i$ doses of IPTp |
| $\boldsymbol{T}_{\boldsymbol{i}}$ | Treated pregnant women who had clinical malaria with $i$ doses of IPTp |
| $\boldsymbol{R}_{\boldsymbol{i}}$ | Recovered pregnant women with $i$ doses of IPTp |
| $\boldsymbol{P}_{\boldsymbol{i,R}}$ | Population with protection of i dose(s) who had a previous malaria infection |
| $\boldsymbol{P}_{\boldsymbol{i,S}}$ | Population with protection of i dose(s) who have no history of malaria infection |

The model equations for the pregnant women who have received zero doses of IPTp are outlined in the equations (1) - (8).

$\frac{dS`}{dt}= \alpha R`- \lambda S`-\theta_{0}S`$	(1)

$\frac{dE`}{dt} = \lambda S` - \sigma_{j}E`- \theta_{0}E`$	(2)

$\frac{dA`}{dt}=P_{i}\sigma_{j}E`+ \omega C`- \delta A`- \theta_{0}A`$	(3)

$\frac{dC`}{dt}=\left( 1-P_{i} \right)\sigma j E`+\epsilon{C`}_{sev0}-\tau C`-\eta_{c}C`-\omega C`- \theta_{0}C`$	(4)

$\frac{d{C`}_{sev0}}{dt}=\eta_{c}C`-\epsilon{C`}_{sev0}-\tau_{sev}{C`}_{sev0}- \mu_{sev}{C`}_{sev0}$	(5)

$\frac{dT`}{dt}=\tau C`- r_{j}T`$	(6)

$\frac{{dT`}_{sev0}}{dt}= \tau_{sev}{C`}_{sev0}- r_{sev}{T`}_{sev0}$	(7)

$\frac{dR`}{dt}=r_{j}T`+\delta A`+ r_{sev}{T`}_{sev0}- \theta_{0}R`-\alpha R`$	(8)

The equations for the first dose of IPTp are outlined by the equations (9) – (18).

$\frac{dP_{1,R}}{dt}=\theta_{0}R`+ \theta_{0}C`+\theta_{0}A`+\theta_{0}E`- \gamma_{1}P_{1,R}-\theta_{1}P_{1,R}$	(9)

$\frac{dP_{1,S}}{dt}= \theta_{0}S`-\rho_{1}P_{1,S}-\theta_{1}P_{1,S}$	(10)

$\frac{dS_{1}}{dt}=\rho_{1}P_{1,S}+\alpha_{1}R_{1}- \lambda S_{1}-\theta_{1}S_{1}$	(11)

$\frac{dE_{1}}{dt}= \lambda S_{1}-\sigma_{j}E_{1}-\theta_{1}E_{1}$	(12)

$\frac{dA_{1}}{dt}={P_{i}\sigma}_{j}E_{1}+ \omega C_{1}-\delta A_{1}-\theta_{1}A_{1}$	(13)

$\frac{dC_{1}}{dt}= {(1-P_{i})\sigma}_{j}E_{1}+ \epsilon C_{sev1}-\tau C_{1}-\eta_{c}C_{1}-\omega C_{1}-\theta_{1}C_{1}$	(14)

$\frac{dC_{sev1}}{dt}=\eta_{c}C_{1}-\epsilon C_{sev1}-\tau_{sev}C_{sev1}-\mu_{sev}C_{sev1}$	(15)

$\frac{dT_{1}}{dt}=\tau C_{1}-r_{j}T_{1}$	(16)
$\frac{{dT}_{sev1}}{dt}= \tau_{sev}C_{sev1}- r_{sev}T_{sev1}$	(17)

$\frac{dR_{1}}{dt}=r_{j}T_{1}+\delta A_{1}+\gamma_{1}P_{1,R}+r_{sev}T_{sev1}-\theta_{1}R_{1}-\alpha_{1}R_{1}$	(18)

The equations for the second dose of IPTp are outlined in the equations (19) – (28).

$\frac{dP_{2,R}}{dt}={\theta_{1}P_{1,R}+\theta}_{1}R_{1}+ \theta_{1}C_{1}+\theta_{1}A_{1}+\theta_{1}E_{1}- \gamma_{2}P_{2,R}-\theta_{2}P_{2,R}$	(19)

$\frac{dP_{2,S}}{dt}= {\theta_{1}P_{1,S}+\theta}_{1}S_{1}-\rho_{2}P_{2,S}-\theta_{2}P_{2,S}$	(20)

$\frac{dS_{2}}{dt}=\rho_{2}P_{2,S}+\alpha_{2}R_{2}- \lambda S_{2}-\theta_{2}S_{2}$	(21)

$\frac{dE_{2}}{dt}= \lambda S_{2}-\sigma_{j}E_{2}-\theta_{2}E_{2}$	(22)

$\frac{dA_{2}}{dt}={P_{i}\sigma}_{j}E_{2}+ \omega C_{2}-\delta A_{2}-\theta_{2}A_{2}$	(23)

$\frac{dC_{2}}{dt}= {(1-P_{i})\sigma}_{j}E_{2}+ \epsilon C_{sev2}-\tau C_{2}-\eta_{c}C_{2}-\omega C_{2}-\theta_{2}C_{2}$	(24)

$\frac{dC_{sev2}}{dt}=\eta_{c}C_{2}-\epsilon C_{sev2}-\tau_{sev}C_{sev2}-\mu_{sev}C_{sev2}$	(25)

$\frac{dT_{2}}{dt}=\tau C_{2}-r_{j}T_{2}$	(26)

$\frac{{dT}_{sev2}}{dt}= \tau_{sev}C_{sev2}- r_{sev}T_{sev2}$	(27)

$\frac{dR_{2}}{dt}=r_{j}T_{2}+\delta A_{2}+ r_{sev}T_{sev2}+\gamma_{2}P_{2,R}-\theta_{2}R_{2}-\alpha_{2}R_{2}$	(28)

The equations for the third dose of IPTp are outlined by the equations (29) – (38).

$\frac{dP_{3,R}}{dt}=\theta_{2}P_{2,R}+\theta_{2}R_{2}+ \theta_{2}C_{2}+\theta_{2}A_{2}+\theta_{2}E_{2}- \gamma_{3}P_{3,R}$	(29)

$\frac{dP_{3,S}}{dt}= \theta_{2}P_{2,S}+\theta_{2}S_{2}-\rho_{3}P_{3,S}$	(30)

$\frac{dS_{3}}{dt}=\rho_{3}P_{3,S}+\alpha_{3}R_{3}- \lambda S_{3}$	(31)

$\frac{dE_{3}}{dt}= \lambda S_{3}-\sigma_{j}E_{3}$	(32)

$\frac{dA_{3}}{dt}={P_{i}\sigma}_{j}E_{3}+ \omega C_{3}-\delta A_{3}$	(33)

$\frac{dC_{3}}{dt}= {(1-P_{i})\sigma}_{j}E_{3}+ \epsilon C_{sev3}-\tau C_{3}-\eta_{c}C_{3}-\omega C_{3}$	(34)

$\frac{dC_{sev3}}{dt}=\eta_{c}C_{3}-\epsilon C_{sev3}-\tau_{sev}C_{sev3}-\mu_{sev}C_{sev3}$	(35)

$\frac{dT_{3}}{dt}=\tau C_{3}-r_{j}T_{3}$	(36)

$\frac{{dT}_{sev3}}{dt}= \tau_{sev}C_{sev3}- r_{sev}T_{sev3}$	(37)

$\frac{dR_{3}}{dt}=r_{j}T_{3}+\delta A_{3}+ r_{sev}T_{sev3}+\gamma_{3}P_{3,R}-\alpha_{3}R_{3}$	(38)

The parameters for the model of IPTp are fully described in Table S2.

**Table S2. Model Parameters.**

| Parameter | Description | Value | Range | Source |
| --- | --- | --- | --- | --- |
| α | Loss of immunity | 1/365 |  | [1] |
| a | Human feeding rate per mosquito | 0.3 | (0.10 – 1.0) | [2] |
| b | Transmission efficiency from mosquitoes to humans | 0.2 | (0.010–0.27) | [2] |
| c | Transmission efficiency from humans to mosquitoes | 0.3 | (0.072–0.64) | [2] |
| $\boldsymbol{\gamma}_{\boldsymbol{m}}$ | Rate of onset of infectiousness in mosquitoes | 1/10 | (5,15) | [3] |
| $\boldsymbol{\mu}_{\boldsymbol{m}}$ | Natural birth/death rate in mosquitoes | 1/15 | (0.012- 0.166) | [2] |
| $\boldsymbol{P}_{\boldsymbol{i}}$ | Proportion of new infections that become asymptomatic among pregnant women | 0.261 | (0.17- 0.37) | [4] |
| $\boldsymbol{\sigma}_{\boldsymbol{j}}$ | Rate at which infected individuals progress through incubation period | 10 days | (9-30) | [5] |
| τ | Treatment seeking rate of clinically ill individuals | 1/4 ${day}^{-1}$ | (0.01- 0.727) | [6] |
| $\boldsymbol{\tau}_{\boldsymbol{sev}}$ | Treatment seeking rate of severely ill individuals | 1/2 ${day}^{-1}$ | (0.03, 1) | Expert opinion |
| $\boldsymbol{\gamma}_{\boldsymbol{i}}$ | Loss of IPTp prophylaxis for the pregnant women with history of i dose who previously had an infection | 42 days | (4-42) | [7, 8] |
| $\boldsymbol{\rho}_{\boldsymbol{i}}$ | Loss of IPTp prophylaxis for the pregnant women with history of i dose but without history of an infection | 42 days | (4-42) | [7, 8] |
| $\boldsymbol{r}_{\boldsymbol{j}}$ | Recovery rate of individuals after malaria treatment | 1/28 | (0.01- 0.08) | [9, 10] |
| δ | Rate of natural recovery of asymptomatic individuals | 1/ (0.5*365) | (0.002- 0.01) | [10] |
| $\boldsymbol{\zeta}_{\boldsymbol{a}}$ | Relative infectiousness of asymptomatic infections | 0.2 | (0-0.50) | [11] |
| $\boldsymbol{\zeta}_{\boldsymbol{t}}$ | Relative infectiousness of treated infections | 0.04 | (0, 0.25) | [12] |
| IPTcov1 | Coverage of IPTp1 | 0.64 | - | [13] |
| IPTcov2 | Coverage of IPTp2 | 0.54 | - | [13] |
| IPTcov3 | Coverage of IPTp3 | 0.42 | - | [13] |
| $\boldsymbol{\eta}_{\boldsymbol{c}}$ | Rate of progressing to severe infection | 6 days | - | Assumed |
| ε | Rate of loss of severe symptoms in untreated severe infections | 1/5 | (1,10) | [12] |
| ω | Rate of loss of symptoms in untreated clinical infection | 1/5 | (5,15) | [12] |
| $\boldsymbol{\mu}_{\boldsymbol{sev}}$ | Rate of death in severe illness | 0.0004 | (0.0001-0.0008) | [14] |
| seasonality amplitude | Amplitude of seasonal variation | 0.89 | (0-1) | Data |
| peak | Month of peak transmission | 1 | - | Data |
| $\boldsymbol{q}_{\boldsymbol{1}}$ | Placental malaria LBW risk | 0.03 | - | [15] |
| $\boldsymbol{q}_{\boldsymbol{2}}$ | Probability of stillbirth | 0.016 | - | [15] |
| w | LBW risk with zero IPTp dose | 0.32 | - | [16] |
| x | LBW risk with one IPTp dose | 0.25 | - | Assumed |
| y | LBW risk with two IPTp doses | 0.17 | - | Assumed |
| z | LBW risk with three IPTp doses | 0.069 | - | [17] |

**Force of infection**

The FOI used in our model is calculated from the simple host-vector SEIR-SEI malaria model given by the equations below:

$\frac{{dS}_{m}}{dt}= \mu_{m}M-ac \frac{I}{P} S_{m}- \mu_{m}S_{m}$	(39)

$\frac{{dE}_{m}}{dt}= ac \frac{I}{P} S_{m}-\left( \gamma_{m}+ \mu_{m} \right)E_{m}$	(40)

$\frac{{dI}_{m}}{dt}= \gamma_{m} E_{m}- \mu_{m} I_{m}$	(41)

$\frac{dS}{dt}= \mu_{h}P-a \frac{M}{P}b\frac{I_{m}}{M}S+ \rho R-\mu_{h}S$	(42)

$\frac{dE}{dt}= a \frac{M}{P}b\frac{I_{m}}{M}S-(\gamma_{h}+ \mu_{h})E$	(43)

$\frac{dI}{dt}= \gamma_{h}E-(r+ \mu_{h})I$	(44)

$\frac{dR}{dt}= rI-(\rho+ \mu_{h})R$	(45)

Where $M= S_{m}+ E_{m}+ I_{m}$	and $P=S+E+I+R$.

Assuming that there is no change in the mosquito population and setting all the vector equations to zero. We let $\lambda_{m}=ac\frac{I}{P}$ be the mosquito FOI and $\lambda_{h}=a \frac{M}{P} b\frac{I_{m}}{M}=ab\frac{I_{m}}{P}$ be the human FOI, substituting and solving equations 39 and 40 for $\lambda_{h}$ while also assuming that $m= \frac{M}{P}$ gives the FOI for the static model as:

$$\lambda_{h}=\frac{(a^{2}bcm\frac{I}{TotalP})}{\left( ac\frac{I}{TotalP}+\mu_{m} \right)*(\frac{\gamma_{m}}{\gamma_{m}+\mu_{m}})}$$

In this study we simulated a mathematical model for a wider population at risk in a high and moderate transmission malaria area for ten years. We included calculations of how the ITN intervention and seasonality alters the force of infection (FOI). We then used the FOI from the wider population model for the pregnant women in our IPTp model.

$$\lambda_{pop}=\left( 1-itn \right)*seas*\frac{(a^{2}bcm\frac{I}{TotalP})}{\left( ac\frac{I}{TotalP}+\mu_{m} \right)*(\frac{\gamma_{m}}{\gamma_{m}+\mu_{m}})}$$

where, $I$ are the infectious compartments which contribute to the spread of the infection,

TotalP is the total population of the wider population, $itn$ represents the bed nets effects and is given by:

$itn=min(ITN,1)*itn\_use*itn\_eff$.

We used the cosine function to incorporate seasonality in the model. The parameters included in the seasonality function were estimated from historical rainfall data for a period of ten years. The seasonality function that we used in the model is given by:

$seas=1+ amp* {(\cos\left( 2\pi*\left( \frac{t}{365}-\emptyset\right) \right))}^{peak}$,

Where, *t* is the time step, *amp* is seasonality amplitude, $\emptyset$ is seasonality phase and these parameters are described in Table S2.

**Estimation of LBW risk**

To estimate the number of deliveries that would result in neonates with LBWs from the simulated cohorts, we used information from literature and adapted an approach used in the mathematical model by Ndaïrou  et al. (2018) [18]. We consider that not every woman who becomes infected with malaria would give birth to a LBW neonate. If the infection sequesters to the placenta it leads to placental infection. Placental infection increases the risk of the pregnant women delivering a LBW neonate [16]. Notably, there is a chance that some women may have a stillbirth, and we included that in our estimation. Receiving IPTp doses protects the women and improves the birth weight of the neonate [19, 20]. The prevalence of malaria related LBW risk differs depending on the number of IPTp doses that a pregnant woman received and we included that in our estimation. However, we excluded the gravidity of the women. The formula that we used to estimate the number of neonates with LBW is shown below:

$$Number of neonates with LBWs=Placental malaria LBW risk*\left( 1-probabibility of stillbirth \right)*\left[ (LBW risk rate with zero IPTp dose*Total number of pregnancies which had infection and zero dose)+( LBW risk rate with one IPTp dose*Total number of pregnancies which had infection and one dose)+(LBW risk with two IPTp doses*Total number of pregnancies which had infection and two doses)+\left( LBW risk with three IPTp doses*Total number of pregnancies which had infection and three doses \right) \right]$$

**Uncertainty ranges**

The Latin Hypercube Sampling (LHS) method was used for sampling parameter values from the multidimensional space and estimating the ranges of uncertainty around parameter values. The distributions chosen to represent uncertainty around parameters are shown in Table S3. Gamma distributions were chosen to represent uncertainty around strictly positive parameter values. The Log normal distribution was used for proportions with skewness. Finally, triangular distribution was used for parameters with known maximum and minimum values.

**Table S3. Probability distributions reflecting uncertainty in parameters.**

| **Parameter** | **Distribution** | **Mean** |
| --- | --- | --- |
| Proportion of new infections that become asymptomatic among pregnant women | Lognormal (0.17, 0.37) | 0.261 |
| Recovery rate of individuals after malaria treatment | Gamma (0.01, 0.08) | 1/28 |
| Rate of death in severe illness | Gamma (0.0001,0.0008) | 0.0004 |
| Rate at which infected individuals progress through incubation period | Gamma (0.04, 0.196) | 1/10 |
| Rate of onset of infectiousness in mosquitoes | Triangular (0.05-0.25) | 1/10 |

**Sensitivity analysis**

We first performed one at a time sensitivity analysis of selected parameters to analyze how changes in each parameter affect the malaria incidence. This involved conducting 50 stochastic simulations of one parameter each time while other parameters remained unchanged, that were sampled from their various probability distributions. For each run, we recorded the cumulative incidence of malaria cases at the end of the simulation and stored this value alongside the sampled value of the parameters. Lastly, we analysed the data to determine the range`s extreme (highest and lowest) parameter values and the associated total cumulative incidence outcomes. We then constructed a tornado plot for these extreme parameter values against the total cumulative incidence and interpreted the results of the plot. Additionally, we conducted multivariate sensitivity analysis, generating ranges for almost all the parameters in our model. We then created random samples of several parameters from their various probability distributions and ran the model in R. Our model results indicate that the most sensitive parameters are **b, m,** and **rho** in that order. The sensitivity analysis and the corresponding values of the parameters are illustrated in Fig S3 and Table S4 respectively. The standardized beta coefficients for the anticipated parameters were then determined using multiple regression, and the results are displayed in Table S5.

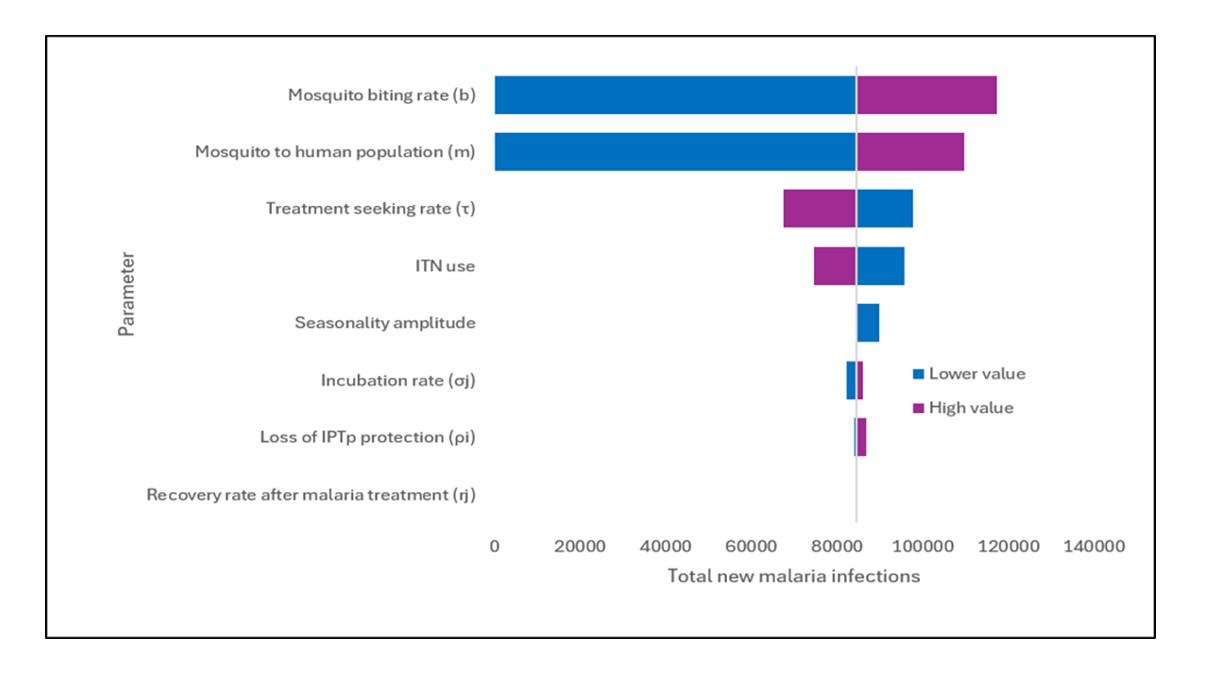

**Fig S3. Sensitivity analysis tornado plot for selected parameters in the IPTp model.** The baseline for the total cumulative incidence is 84,534 and is shown by the vertical line between 80,000 and 100,000.

**Table S4. Baseline values and sensitivity values for parameters for the IPTp model.**

| **Parameter** | **High value** | **Base case** | **Lower value** |
| --- | --- | --- | --- |
| ITN use | 96% | 70% | 32% |
| Mosquito to human population ratio | 4.96 | 3 | 1.04 |
| Seasonality amplitude | 0.99 | 0.89 | 0.01 |
| Mosquito biting rate | 0.468 | 0.3 | 0.21 |
| Loss of IPTp protection | 0.18 (6 days) | 0.02 (42days) | 0.01667 (60days) |
| Recovery rate after malaria treatment | 0.056 (18 days) | 1/28 (28 days) | 0.016 (62.5 days) |
| Incubation rate | 0.3 (3 days) | 0.1 (10 days) | 0.05 (20 days) |
| Treatment seeking rate | 0.446 (2 days) | 0.25 (4 days) | 0.05 (20 days) |

**More results**

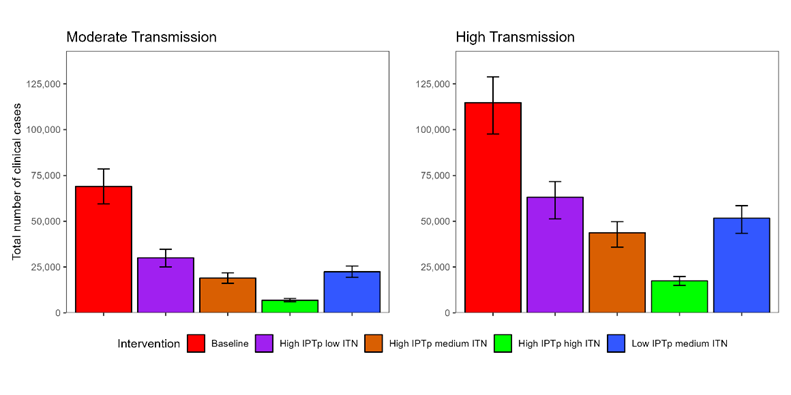

**Fig S4. Total clinical cases for pregnancies that started in August under different intervention scenarios.** Bars show the average total number of clinical cases and their respective 90% uncertainty interval derived from parameter variability.

**Table S5. Total clinical cases under different combinations of interventions in moderate and high transmission setting at the end of the ten-month pregnancy period for pregnancies that started in August.** The uncertainty interval for the total number of clinical cases is presented in parenthesis next to the corresponding average total number of clinical cases.

| **Scenario** | **Total clinical cases**  **Transmission setting** | |
| --- | --- | --- |
|  | **Moderate** | **High** |
| Baseline: | 69,024  (59,474 – 78,535) | 114,749  (97,686 – 128,891) |
| High IPTp low ITN | 30,056  (25,095 – 34,699) | 63,128 (51,305 – 71,706) |
| High IPTp medium ITN | 18,989  (16,109 – 21,857) | 43,761  (35,881 – 49,771) |
| High IPTp high ITN | 6,886  (6,000 – 7,805) | 17,450  (14,932 – 19,797) |
| Low IPTp medium ITN | 22,406  (19,340 – 25,585) | 51,659  (43,415 – 58,524) |

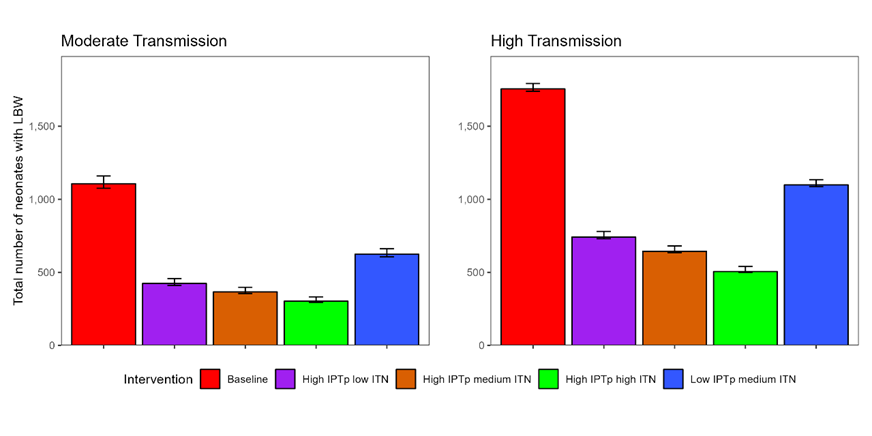

**Fig S5. Total number of neonates with LBW for pregnancies that started in August under different intervention scenarios.** Bars show the average number of neonates with LBW and their respective 90% uncertainty interval derived from parameter variability.

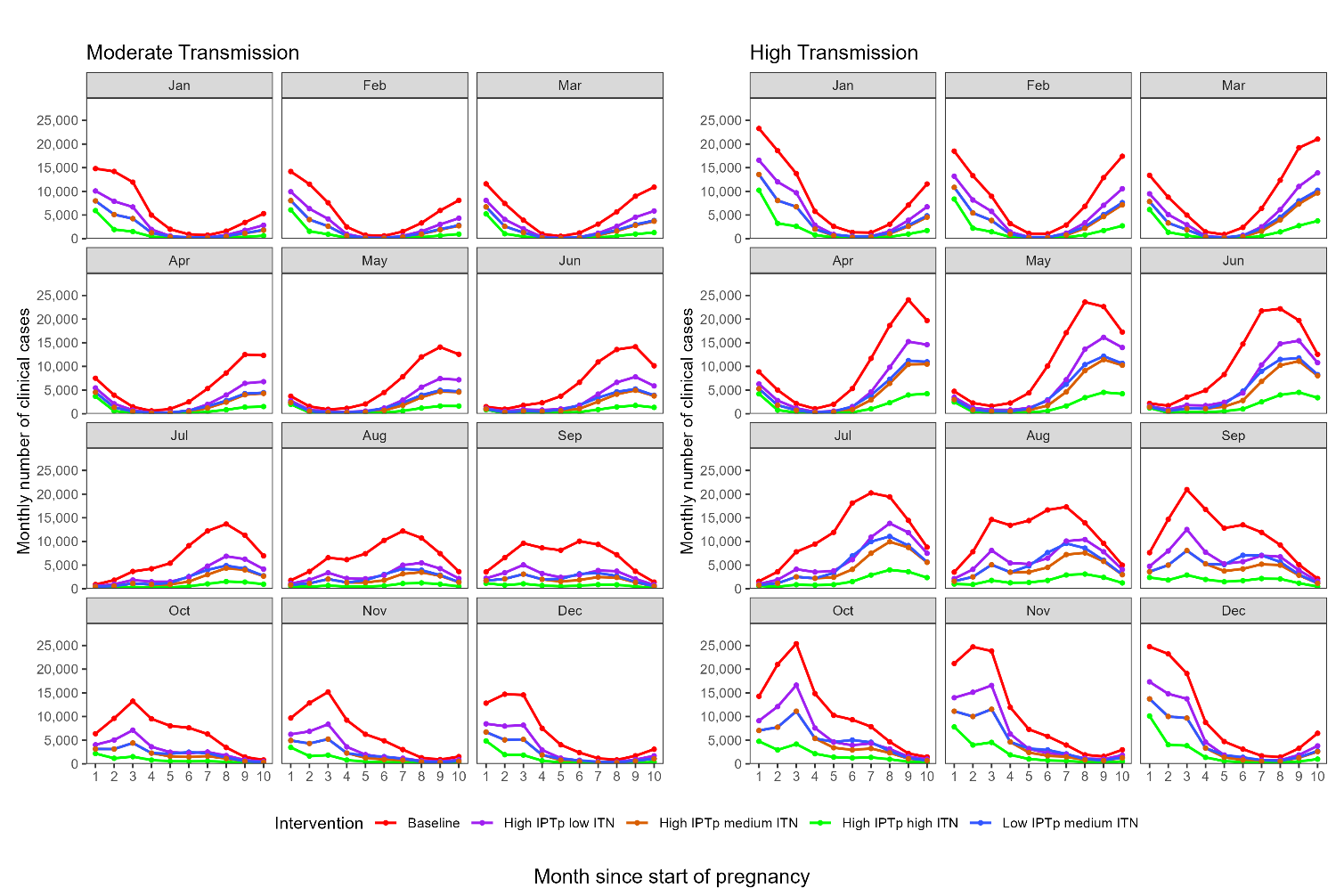

**Fig S6. Comparison of the monthly number of clinical cases in high and moderate transmission setting under intervention scenarios for different pregnancy cohorts with different start times.** The months highlighted in grey rectangular boxes represent the different months of start of pregnancies.

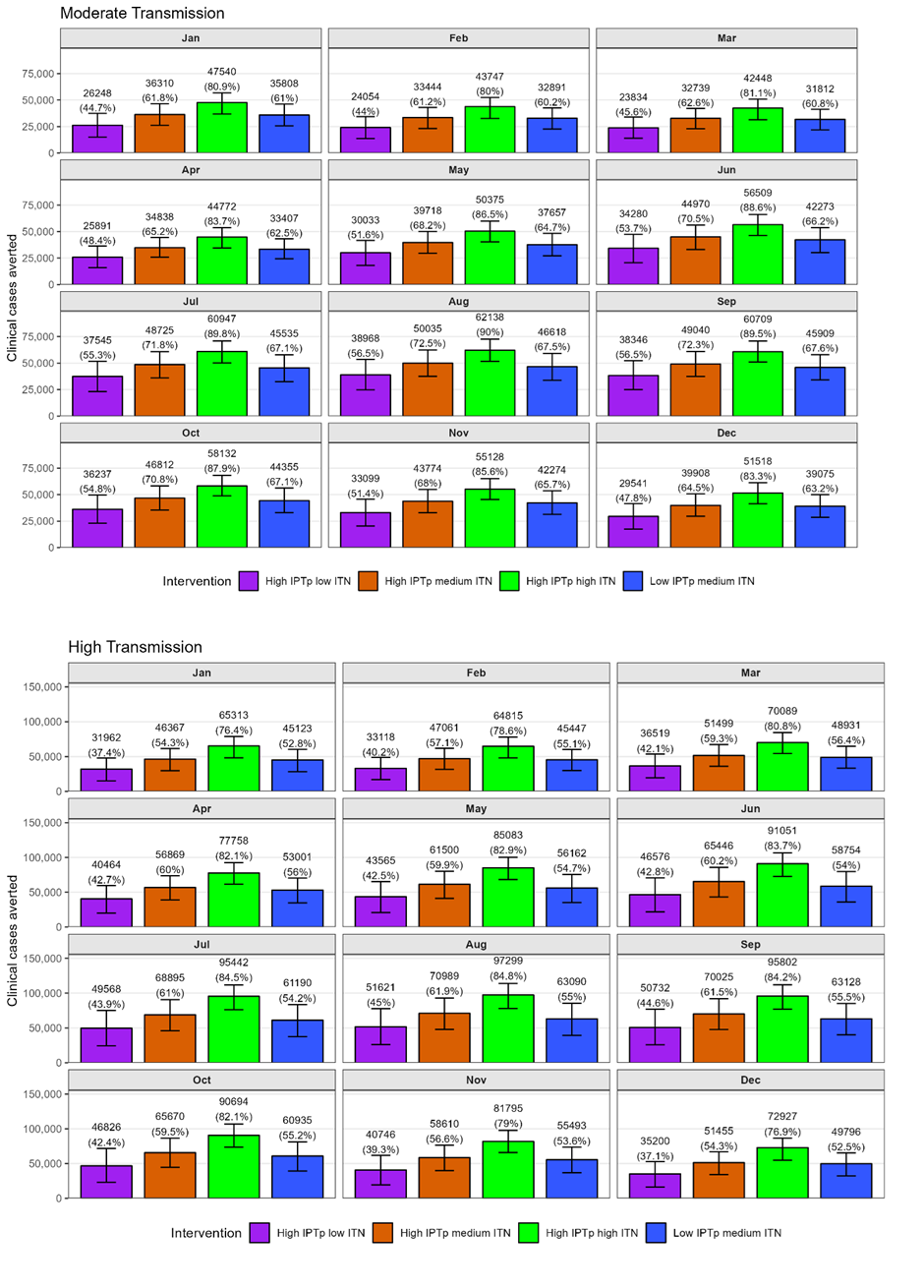

**Fig S7. Number and percentages of clinical cases averted under the different intervention scenarios described in Table 1 in moderate and high transmission.** Bars show the average number of clinical cases averted and their respective 90% uncertainty interval derived from parameter variability. The months highlighted in grey rectangular boxes represent the different months of start of pregnancies.

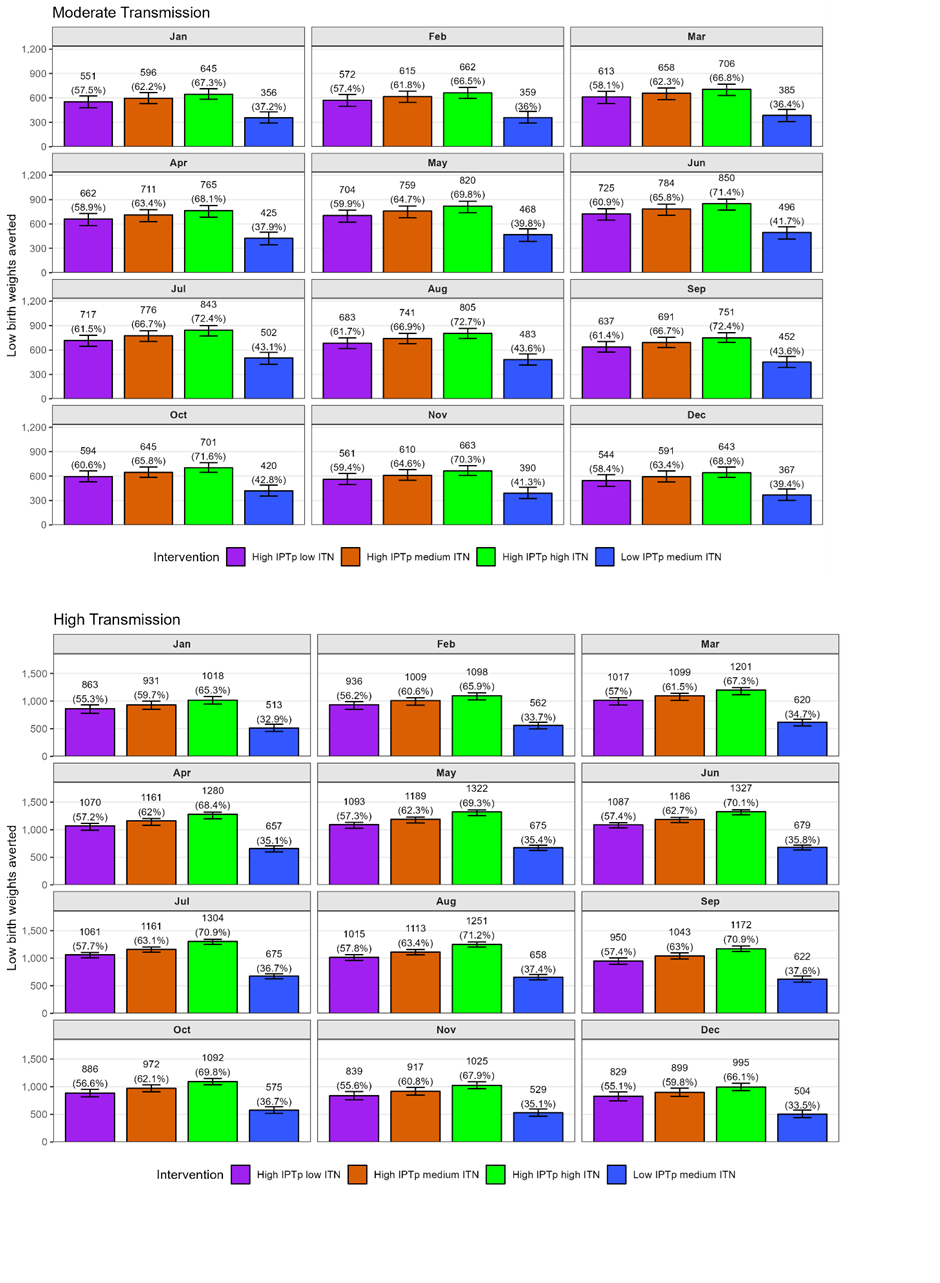

**Fig S8. Percentages of Low birth weight averted under the different intervention scenarios relative to the baseline in moderate and high transmission.** Bars show the number of LBW averted and their respective 90% uncertainty interval derived from parameter variability. The months highlighted in grey rectangular boxes represent the different months of start of pregnancies.

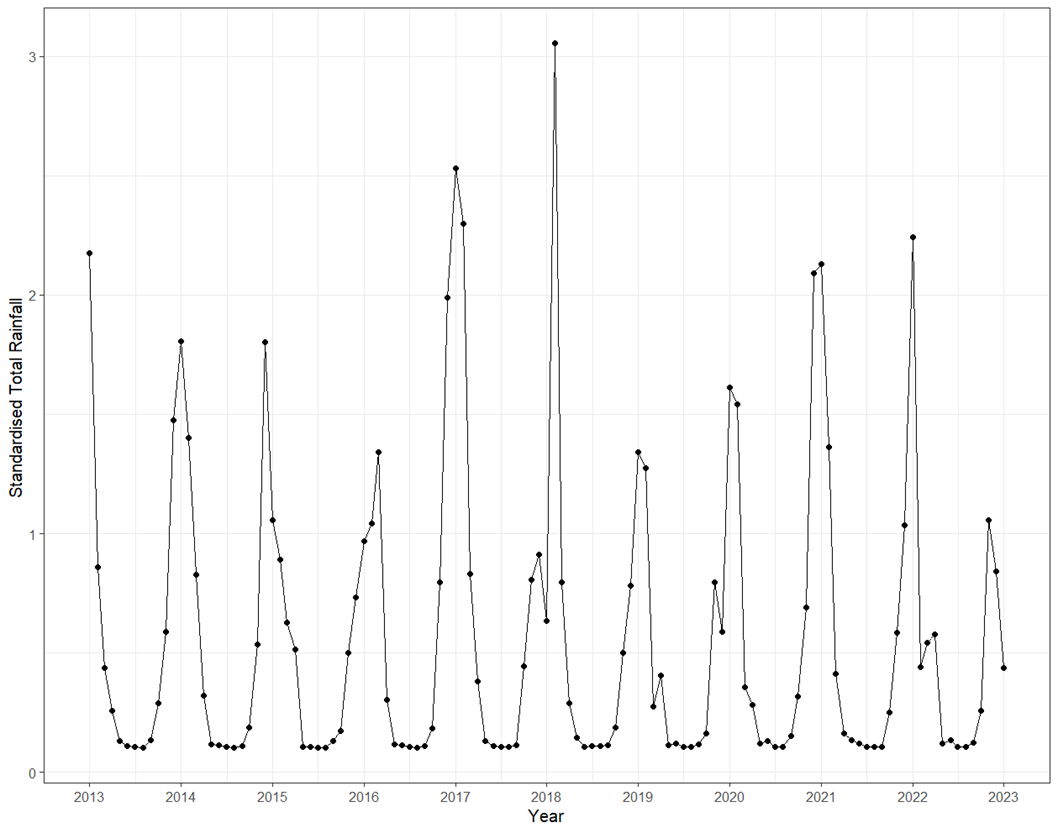

**Fig S9. Standardised total monthly rainfall for Zimbabwe for a period of 10 years from 2013 to 2023.** This data was used to parameterise the mathematical model.

**Table S5. Standardised Beta coefficients and multiple regression results for sensitivity analysis.**

| **Multiple regression estimation** | | |  | **Standardised and ordered coefficients** | |
| --- | --- | --- | --- | --- | --- |
| **Parameter** | **estimate** | **P-value** |  | **Parameter** | **Beta coefficient** |
| b | 419424.77 | <0.001 |  | **b** | **0.599** |
| ρ | 14403322.25 | <0.001 |  | **ρ** | **0.526** |
| m | 17112.57 | <0.001 |  | **m** | **0.376** |
| $r_{a}$ | -3030102.9 | <0.001 |  | $\sigma_{j}$ | 0.043 |
| ITN efficacy | -34279.05 | 0.015 |  | $\eta_{c}$ | 0.024 |
| $\tau_{sev}$ | -6074.5 | 0.045 |  | $\gamma_{1}$ | 0.012 |
| $\sigma_{j}$ | 52179.78 | 0.17 |  | ω | 0.0058 |
| $\eta_{c}$ | 1085.68 | 0.44 |  | $r_{j}$ | 0.002 |
| τ | -4648.2 | 0.48 |  | $r_{a}$ | -0.173 |
| ITN use | -5760.5 | 0.6 |  | ITN efficacy | -0.075 |
| $\gamma_{1}$ | 140.71 | 0.69 |  | $\tau_{sev}$ | -0.062 |
| $P_{i}$ | -10358.0 | 0.74 |  | τ | -0.022 |
| ε | -139.9 | 0.83 |  | ITN use | -0.016 |
| ω | 105.86 | 0.85 |  | $P_{i}$ | -0.010 |
| $r_{c}$ | -9701.86 | 0.91 |  | ε | -0.0069 |
| $r_{j}$ | 5807.07 | 0.94 |  | $r_{c}$ | -0.0036 |
| $\mu_{sev}$ | -172629.9 | 0.98 |  | $\mu_{sev}$ | -0.0007 |
| (Intercept) | -48185.8664 | 0.011 |  |  |  |
